# Exploring multidrug resistance patterns in community-acquired *E. coli* urinary tract infections with machine learning

**DOI:** 10.1101/2025.02.05.25321745

**Authors:** Elise Hodbert, Olivier Lemenand, Sonia Thibaut, Thomas Coeffic, David Boutoille, Stephane Corvec, Gabriel Birgand, Laura Temime, French Clinical Laboratories Nationwide Network

## Abstract

**Background:** While associations of antibiotic resistance traits are not random in multidrug-resistant (MDR) bacteria, clinically relevant resistance patterns remain relatively underexplored. This study used machine learning, specifically association-set mining, to explore resistance associations within *E. coli* isolates from community-acquired urinary tract infections (UTIs).

**Methods:** We analysed antibiograms of community-acquired *E. coli* UTI isolates collected from 2018 to 2022 by France’s national surveillance system. Association-set mining was applied separately to extended-spectrum beta-lactamase-producing *E. coli* (ESBL-EC) and non-ESBL-EC. MDR patterns that had expected support (reflecting pattern frequency) and conditional lift (reflecting association strength) higher than expected by chance (p-value≤0.05) were used to construct resistance networks, and analysed according to time, age and gender.

**Findings:** The number of isolates increased from 360 287 in 2018 (10 150 ESBL-EC, 350 137 non-ESBL-EC) to 629 017 in 2022 (18 663 ESBL-EC, 610 354 non-ESBL-EC). More MDR patterns were selected in ESBL-EC than non-ESBL-EC (2022: 1770 vs 93 patterns), with higher respective network densities (2022: 0.230 vs 0.074). Fluoroquinolone, third-generation cephalosporin and penicillin resistances were strongly associated in ESBL-EC. The median densities of resistance association networks increased from 2018 to 2022 in both ESBL-EC (0.238 to 0.302, p-value=0.06, Pearson test) and non-ESBL-EC (0.074 to 0.100, p-value=0.04). Across all years, median network densities were higher in men than women in both ESBL-EC (2022: 0.305 vs 0.276) and non-ESBL-EC (2022: 0.128 vs 0.094); they were also higher in individuals over 65 years old than under 65 in ESBL-EC (2022: 0.289 vs 0.275) and non-ESBL-EC (2022: 0.103 vs 0.088).

**Interpretation:** These findings, which show increasing MDR associations, especially in men and older individuals, highlight the importance of ongoing resistance surveillance to understand the future evolution of resistance patterns.

**Funding:** This work received funding from the French government through the National Research Agency project COMBINE ANR–22-PAMR-0003.

**Research in context:** *Evidence before this study:* We searched Pubmed for previously published articles without any date or language restrictions using the search terms (multiresistan* OR “multidrug-resistan*”) AND (“data mining” OR “machine learning” OR “artificial intelligence”) AND (pattern* OR associat*). We found three studies that used machine learning to identify multiresistance patterns in various pathogens (chicken-associated *Escherichia coli*, human-associated *Staphylococcus aureus* and cattle-associated *Salmonella enterica*) in the United States. However, to our knowledge, no machine-learning studies to date have explored multiresistance patterns in human-associated Enterobacterales, especially within European contexts.

*Added value of this study:* Our study provided a novel and detailed analysis of multiresistance patterns in community-acquired *E. coli* urinary tract infection collected from a French national surveillance system. Our findings confirmed that association-set mining is effective for identifying resistance associations in antibiotic resistance surveillance data. We explored the temporal evolution of resistance associations, gender-specific and age-specific differences, which to our knowledge, had not been previously analysed.

*Implications of all the available evidence:* Our results suggest a temporal increase of resistance associations in community-acquired *E. coli* UTI and identify key patterns in different subpopulations. In the context of rising antibiotic resistance, optimizing the use of current medications is crucial, as few new antibiotics have been developed in the past two decades. With further research, this work could provide insight for targeted antibiotic stewardship strategies.

## Introduction

Antimicrobial-resistance (AMR) is an increasing public health issue, notably due to the extensive use of antibiotics and increasing global mobility. Infections caused by multidrug-resistant bacteria are challenging to treat, resulting in increased mortality, prolonged hospital stays and higher healthcare costs (1). Initially regarded as a hospital-specific issue, AMR is now recognized as a major problem in community settings and a one-health issue (2). In 2021, AMR-associated infections were directly responsible for approximately 1.14 million deaths globally (3). Multiple studies forecast an increase in resistance to available antibiotics, which could cause >8 million deaths in 2050 (3). The global burden of AMR is driven by a few leading microorganisms, among which resistant *E. coli* infections caused the highest number of deaths in 2019 (4).

Multidrug resistance (MDR) does not result from individual drug resistances occurring together by chance, as documented in multiple bacteria (5). Beyond selection pressure, several biological mechanisms are known to contribute to MDR emergence, including horizontal gene transfer (6), non-specific mechanisms like efflux pumps and specific mechanisms like beta-lactamases (5).

Here, we use association-set mining, a method that has proven effective in identifying patterns of antibiotic resistance traits in chicken-associated *Escherichia coli*, human-associated *Staphylococcus aureus* and cattle-associated *Salmonella enterica* (7–9), to identify multiresistance patterns in community-acquired *E. coli* urinary tract infections (UTIs) collected by the French national surveillance system of antibiotic resistance. We create graphical networks from the patterns detected by the algorithm in order to highlight resistance associations of interest. By comprehensively studying MDR patterns, we aim to better understand their evolution over time and potential differences across subpopulations.

## Methods

### Data source

This study used retrospective data collected via PRIMO (Surveillance and Prevention of Antibiotic Resistance in Primary Care and Nursing Homes), a French nationwide surveillance system of clinical laboratories participating on a voluntary basis (742 laboratories in 2018, 1773 laboratories in 2022) (10). Data included antibiotic susceptibility testing (AST) results, patient’s age, gender, and administrative region. We restricted the analyses to AST collected from *E. coli* UTI collected in community laboratories between January 1^st^, 2018 and December 31^st^, 2022.

AST and Extended-spectrum beta-lactamases (ESBL) testing were carried out and assessed according to the recommendations of the AST committee of the French Microbiology Society (11). Each isolate was tested against 3 to 35 antibiotics and categorized in a Susceptible, Susceptible to increased exposure or Resistant format. For this analysis, Susceptible to increased exposure and Resistant were grouped as Resistant. We excluded antibiotics that were tested on less than 10% of isolates on average across all years, and included a single representative antibiotic when several antibiotics had similar resistance patterns. This process resulted in the inclusion of 27 antibiotics (Supplementary Table 1). Carbapenemase-producing isolates were excluded from the analysis as their low frequency did not allow for a robust analysis of their specific multiresistance patterns. To avoid duplicates, when multiple isolates with the same susceptibility pattern were collected from the same individual (same date of birth and sex) within a single clinical laboratory, only the first isolate in the dataset was included.

Because the analysis was performed using anonymized surveillance data, ethical consent was not required according to the French Data Protection Act. The dataset was accredited by the French National Data Protection Commission (CNIL 1,685,003), and the fully anonymized data waiver for informed consent of study participants was applied.

### Data stratification

We classified isolates into two phenotype categories: ESBL-producing *E. coli* (ESBL-EC), either alone or combined with another phenotype, and non-ESBL-producing *E. coli* (non-ESBL-EC). We conducted analyses separately for each year and phenotype dataset. To explore potential gender- and age-based differences in multiresistance patterns, we also conducted stratified analyses by sex and separate analyses for individuals aged under and over 65.

### Testing the independence of individual resistances

We first assessed the independence of individual resistances (8). In the following, H0 refers to the hypothesis that assumes all resistance traits to antibiotics are mutually independent. We simulated 100 datasets under H0, that matched the observed datasets in size, resistance prevalence, and positions of missing AST results. Resistance traits were generated as independent binomial random variables (n = number of isolates tested; p = resistance prevalence). To evaluate the likelihood of the observed MDR prevalence under H0, we compared the distribution of resistance counts per isolate between observed and simulated data using a Kolmogorov-Smirnov test.

### Multiresistance patterns identification

We identified MDR patterns from our datasets using association-set mining with the *Apriori* algorithm, as previously proposed by Cazer et al. (8,12). Briefly, we first explored the datasets to select all patterns present with a given frequency, setting the required initial minimum frequency at 0.01 for ESBL-EC and 0.001 for non-ESBL-EC. In a second step, we pruned patterns for which two important metrics, expected support (eSup) and conditional lift (cLift), were significantly lower than expected by chance. To do this, we determined the 5^th^ percentile of eSup and cLift under the resistance independence hypothesis H0 using the previously described 100 datasets simulated under H0. eSup measures the pattern frequency within the dataset, while cLift assesses the frequency of a pattern relative to its expected frequency under the assumption that resistance to antibiotics within the pattern are independent. More details on multiresistance pattern identification and accounting for missing data are provided in Supplementary Text 2.

### Construction and analysis of resistance associations networks

We decomposed the MDR patterns remaining after pruning into pairwise associations, and built resistance association networks, where nodes represent antibiotics and edges represent resistance associations between two antibiotics.

For each reconstructed network, we computed network density. Confidence intervals were estimated using: (i) for 2018, 10 datasets generated by a bootstrap procedure of the original dataset, (ii) for 2019 to 2022, 10 random subsamples of the original datasets, each created to match the size of the 2018 dataset. We calculated the 95% confidence intervals from the 2.5^th^ and the 97.5^th^ percentiles of the results. Time changes in network density were assessed using these constant-size yearly samples, and a Pearson trend test.

Networks were also compared between men and women, and individuals under or over 65 years old. Similarly to between-year comparisons, we generated 10 random subsamples of the women’s (resp. over-65) dataset to match the size of the men’s (resp under-65) dataset and computed 95% confidence intervals from either bootstrapped data or these subsamples.

### Sensitivity analysis

The regional contributions to the national dataset varied over the years. To investigate the impact of these variations on our results, we created a dataset where the number of isolates from each region was proportional to its population (details in Supplementary Text 3), and replicated our analyses using this dataset.

All analyses were performed in R 4.3.2, notably using the package “arules” for association-set mining (Hahsler et al., 2018).

## Results

### Data description

The number of isolates increased over time, from 360 287 in 2018 to 628 993 in 2022 (Table 1; p-value=0.01, Pearson trend test). The proportion of ESBL isolates was comprised between 2.8% and 3.0%, with no increasing or decreasing trend across the years (p-value=0.79). Throughout all years, between 84.0% and 84.5% of isolates came from women. The mean age ranged from 58 to 60. The isolates came from all regions of metropolitan France, with varying proportions across the years (Figure S1).

**Table 1:** Summary of PRIMO datasets for *E. coli* UTI samples from 2018 to 2022.

|  | 2018 | 2019 | 2020 | 2021 | 2022 |
| --- | --- | --- | --- | --- | --- |
| **Number of isolates** | 360 287 | 450 820 | 450 135 | 571 126 | 628 993 |
| *ESBL* | 10 150<br>(2.8%) | 13 559<br>(3.0%) | 13 265<br>(2.9%) | 16 006<br>(2.8%) | 18 663<br>(3.0%) |
| *Non-ESBL* | 350 137<br>(97.2%) | 437 261<br>(97.0%) | 436 870<br>(97.1%) | 555 120<br>(97.2%) | 610 330<br>(97.0%) |
| *Women* | 276 220<br>(84.5%) | 356 002<br>(84.4%) | 361 758<br>(84.0%) | 457 789<br>(84.3%) | 520 081<br>(84.1%) |
| *Men* | 50 706<br>(15.5%) | 66 000<br>(15.6%) | 68 730<br>(16.0%) | 85 444<br>(16.7%) | 98 055<br>(15.9%) |
| **Mean age** | 58 | 59 | 59 | 60 | 60 |

### Individual antibiotic resistance prevalence in ESBL-EC and non-ESBL-EC

Regarding penicillins, nearly all ESBL isolates were resistant to ampicillin, amoxicillin and ticarcillin (Figure 1A). Most ESBL isolates were resistant to second- and third-generation cephalosporins, including cefuroxime (100%, 1 535/1 535 tested in 2022), ceftriaxone (99.6%, 18 592/18 663 tested) and cefotaxime (99.6%, 18 592/18 663 tested). For fluoroquinolones, between 70% and 85% of isolates were resistant to ciprofloxacin, levofloxacin, norfloxacin and ofloxacin. However, most ESBL isolates were sensitive to first-line antibiotics including fosfomycin (4.9%, 834/17 180 tested), nitrofurantoin (1.9%, 348/18 259 tested), mecillinam (10.8%, 1 800/16 684 tested) and cefoxitin (11.1%, 2035/18 380 tested).

**Figure 1:**
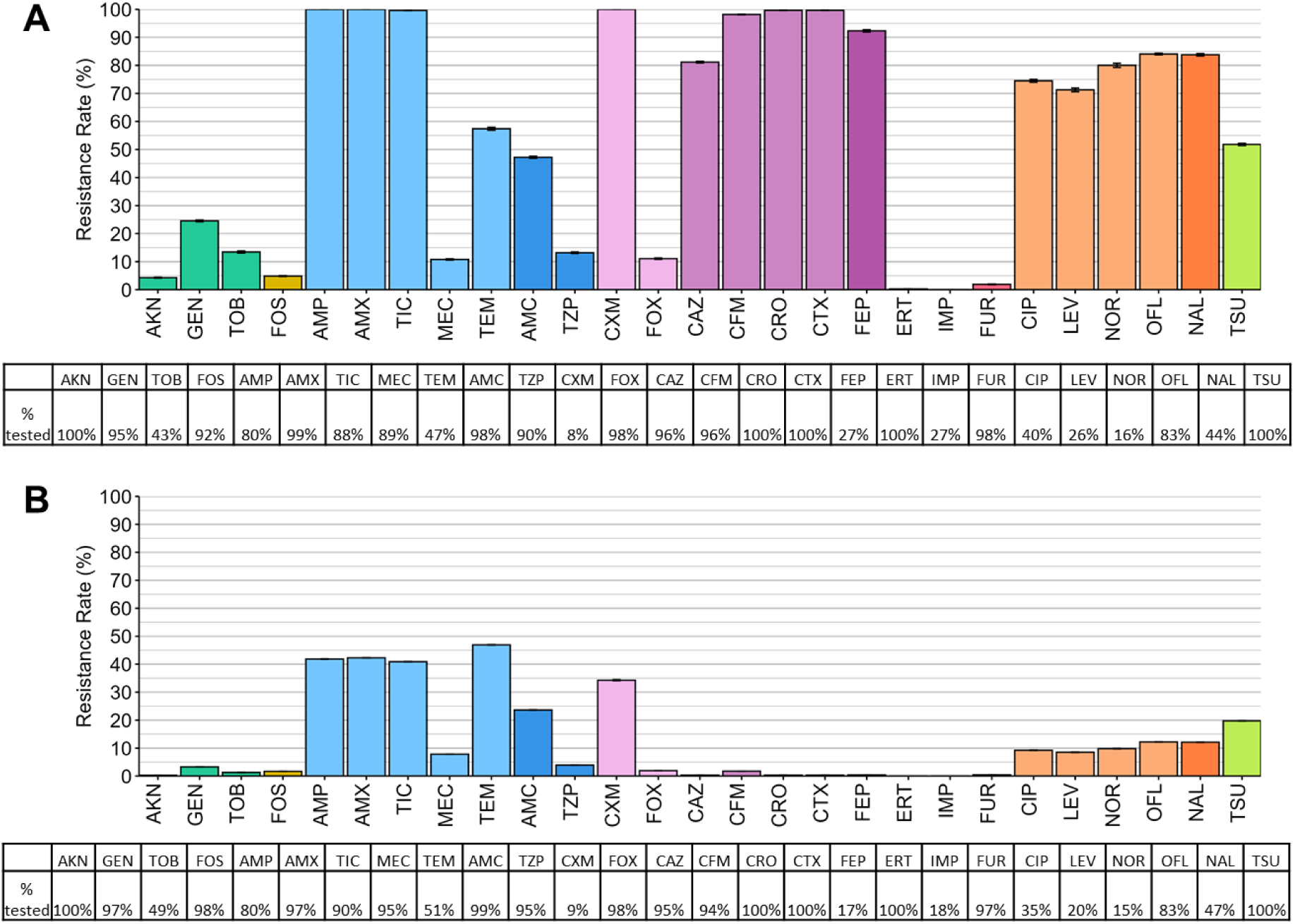
Resistance prevalence and AST frequency for all antibiotics in 2022 in (A) ESBL-EC and (B) non-ESBL-EC. See supplementary Table 1 for a complete list of antibiotics with their full name.

Non-ESBL isolates were often resistant to penicillins including ampicillin (41.8%, 203 547/486 592 tested), amoxicillin (42.3%, 251 107/593 743 tested) and ticarcillin (40.9%, 223 404/546 477 tested) (Figure 1B). Less than 1% of isolates were resistant to third generation cephalosporins including ceftazidime, cefixime, ceftriaxone and cefotaxime. For fluoroquinolones, between 5% and 15% of isolates were resistant to ciprofloxacin, levofloxacin, norfloxacin and ofloxacin. Similar individual antibiotic resistance prevalences were found from 2018 to 2021 (Figure S2).

### Independence of individual resistance traits

The observed distribution of the number of resistances per isolate significantly differed from the simulated distribution under the hypothesis of independence (p-value = 6.81.10^−3^, Kolmogorov-Smirnov test), suggesting that individual resistances were not independent (Figure 2). In the observed dataset, 39% of isolates were pansusceptible, compared to 9% in the simulated dataset. Similar results were obtained when comparing the distributions of the number of resistances per isolate in observed and simulated data from 2018 to 2021 (Figure S3). We also compared the distributions separately for ESBL-EC and non-ESBL-EC from 2018 to 2022 (Figure S4), underlining that the difference was mostly driven by non-ESBL-EC.

**Figure 2:**
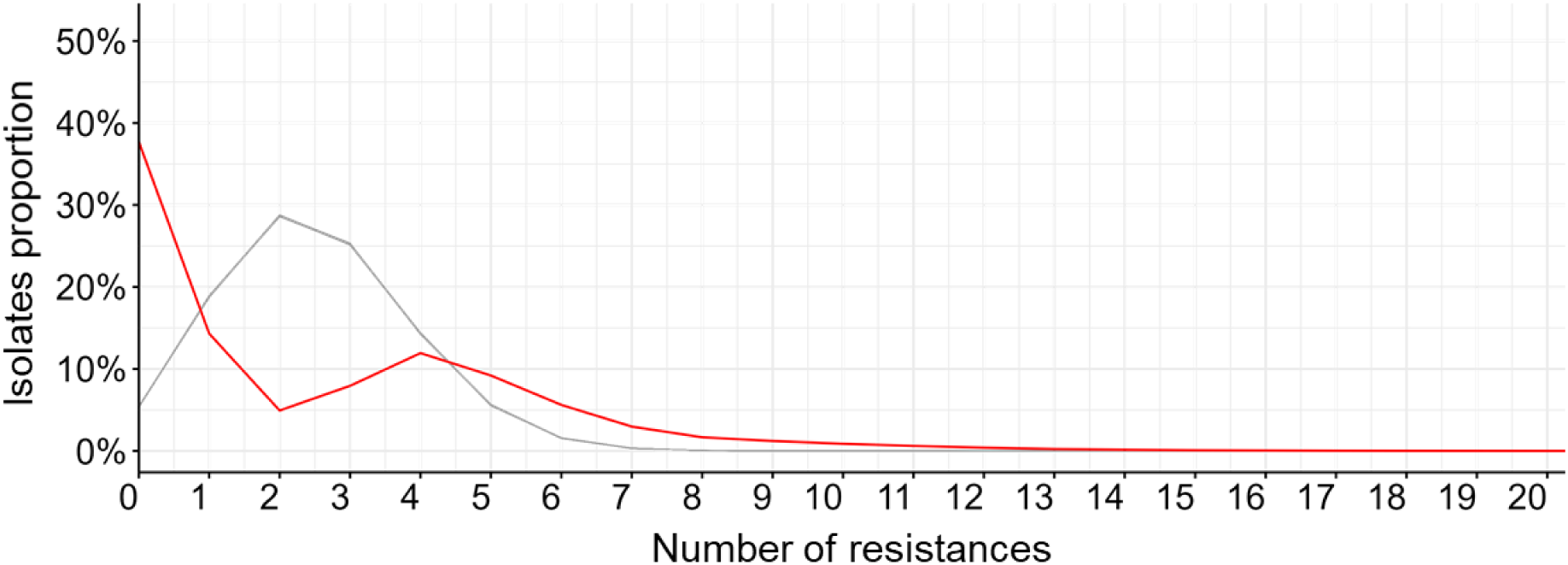
Distribution of the number of resistances per isolate in 2022 for all isolates. The red line represents the observed dataset, and the grey lines represent the simulated datasets.

### Multiresistance pattern selection

For ESBL-EC, the *Apriori* algorithm initially identified 52 824 patterns in 2018 and 123 116 patterns in 2022, with an increasing trend over the years (p-value=0.01, Pearson test) (Table 2). For non-ESBL-EC, the algorithm initially generated 2 529 patterns in 2018 and 4 515 patterns in 2022, with a non-significant increasing trend (p-value=0.06, Pearson test).

**Table 2:** Number of multiresistance patterns selected by association-set mining for each year and phenotype subset. For example, in 2018 ESBL-EC, 52 736 patterns were identified. Cut-off values were set at eSup ≥ 0.257 and cLift ≥ 1.076, meaning that a pattern was retained if it appeared in at least 25.7% of isolates and occurred 7.6% more frequently than expected under the assumption of independence between resistance traits. After pruning, 483 patterns were selected.

| Phenotype | Year | Number of patterns initially | eSup cut-off value | cLift cut-off value | Number of patterns selected |
| --- | --- | --- | --- | --- | --- |

|  |  | generated<br>by the<br>Apriori<br>algorithm |  |  | after<br>pruning |
| --- | --- | --- | --- | --- | --- |
| ESBL | 2018 | 52 736 | 0.257 | 1.076 | 483 |
|  | 2019 | 86 454 | 0.236 | 1.070 | 1 234 |
|  | 2020 | 106 625 | 0.242 | 1.073 | 1 268 |
|  | 2021 | 115 425 | 0.262 | 1.065 | 1 921 |
|  | 2022 | 123 116 | 0.271 | 1.058 | 1 770 |
| Non-ESBL | 2018 | 2 527 | 0.030 | 1.056 | 59 |
|  | 2019 | 3 936 | 0.029 | 1.051 | 82 |
|  | 2020 | 3 756 | 0.030 | 1.052 | 75 |
|  | 2021 | 4 945 | 0.031 | 1.045 | 88 |
|  | 2022 | 4 502 | 0.034 | 1.041 | 93 |

For 2022, the eSup cut-off values were 0.271 for ESBL-EC isolates and 0.034 for non-ESBL-EC, meaning that a pattern was selected if it appeared in at least 27.1% of ESBL-EC or 3.4% of non-ESBL-EC. This value was relatively stable over time for both ESBL (range: 23.6-27.1%, p-value=0.22, Mann-Kendall test) and non-ESBL-EC (range: 2.9-3.4%, p-value=0.13, Mann-Kendall test).

The cLift cut-off value for ESBL-EC and non-ESBL-EC was respectively 1.058 and 1.041 in 2022, meaning that a pattern was selected if it was 5.8% (resp. 4.1%) more frequent than expected by chance under the assumption of resistance independence in ESBL-EC (resp. non-ESBL-EC). These cut-offs were again relatively stable over time for both ESBL-EC (p-value=0.46, Mann-Kendall test) and non-ESBL-EC (p-value=0.09, Mann-Kendall test).

The pruning step selected respectively 1 770 (1.43% of patterns) and 93 (2.06%) patterns for ESBL-EC and non-ESBL-EC in 2022. The number of selected patterns had an increasing trend from 2018 to 2022 for both ESBL-EC (p-value=0.03, Pearson test) and non-ESBL-EC (p-value=0.05, Pearson test).

### Networks of resistance associations

#### Resistance associations networks have higher density for ESBL-EC

Resistance associations networks were much denser in ESBL-EC than non-ESBL-EC, with densities of respectively 0.301 and 0.100 in 2022 (Figure 3). As expected, resistance associations in ESBL-EC involved mainly penicillins, third-generation cephalosporins and fluoroquinolones. In non-ESBL-EC, strong resistance associations were found between penicillins, combinations of penicillin and beta-lactamase inhibitor and quinolones. Regarding first-line antibiotics, almost no significant association involving nitrofurantoin, fosfomycin, mecillinam nor any aminoglycoside was found in both ESBL- and non-ESBL-EC. However, trimethoprim/sulfamethoxazole was associated with third-generation cephalosporins in ESBL-EC and with penicillins in non-ESBL-EC.

**Figure 3:**
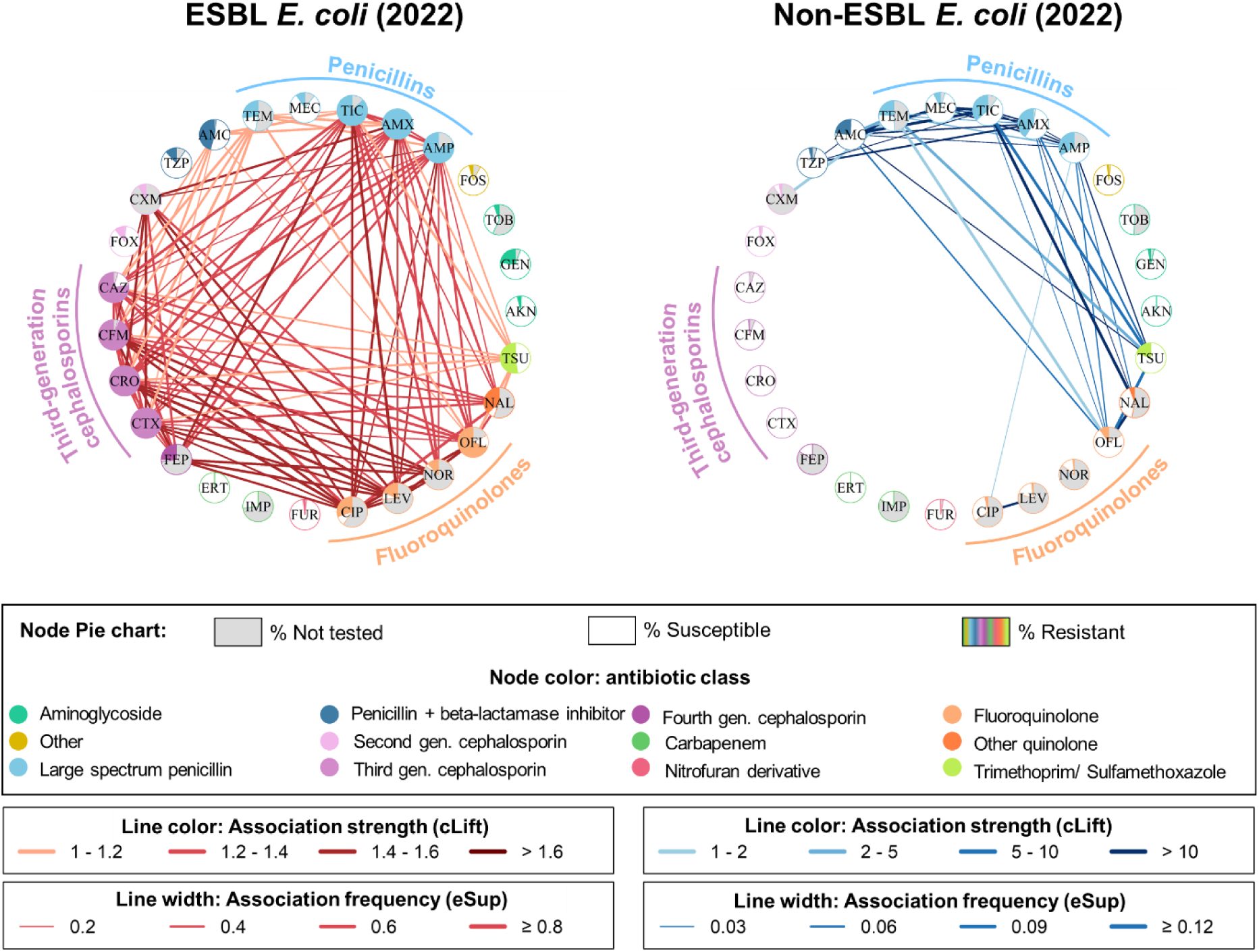
Networks generated from ESBL-EC (A) and non-ESBL-EC (B) in 2022. Patterns were decomposed into nodes (antibiotics) and edges. Nodes are colored with a pie chart: the grey part is the proportion of isolates not tested for this antibiotic, the white part is the proportion of isolates susceptible to this antibiotic and the colored part is the proportion of isolates resistant to this antibiotic. Node color reflects the antibiotic class. Edge color is darker with a high cLift value, meaning that the two antibiotics are substantially more frequently together in a resistance pattern than expected if resistance traits were independent. Edge width is higher with a high eSup value, meaning that the two antibiotics are frequently found together in a resistance pattern. See supplementary Table 1 for a complete list of antibiotics with their full name.

#### Network density increases over time for both ESBL-EC and non-ESBL-EC

Network density increased over time, from 0.238 [0.234; 0.249] in 2018 to 0.301 [0.294; 0.302] in 2022 for ESBL-EC (p-value=0.06, Pearson test), and from 0.074 [0.074; 0.074] in 2018 to 0.100 [0.100; 0.103] in 2022 for non-ESBL-EC (p-value=0.04, Pearson test) (Figure 4).

**Figure 4:**
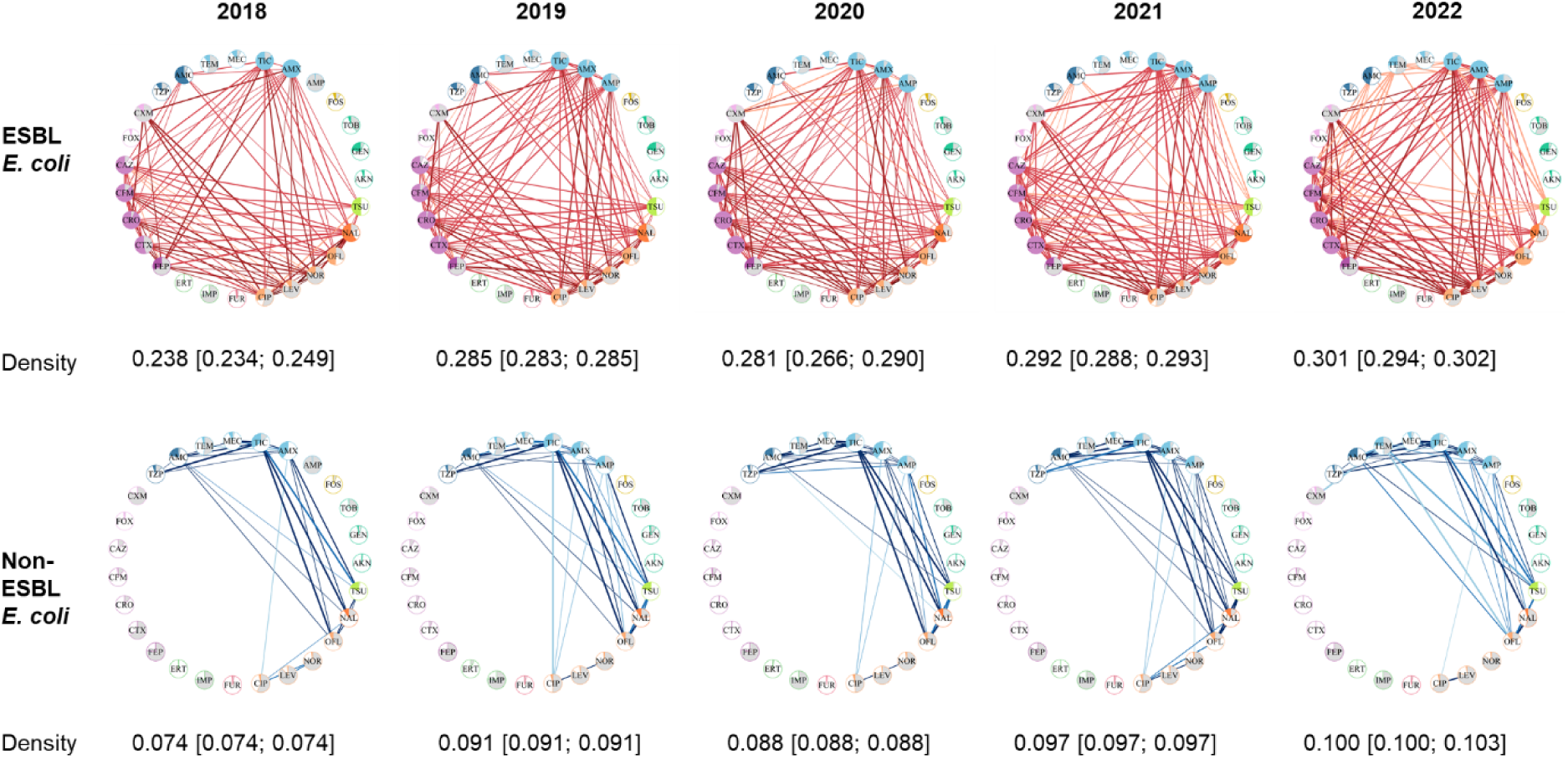
Networks of resistance associations from 2018 to 2022. Median density and 95% confidence interval calculated using: (i) for 2018, 10 bootstrapped datasets, (ii) for 2019 to 2022, 10 subsamples randomly drawn to match the size of the 2018 dataset. See supplementary Table 1 for a complete list of antibiotics with their full name.

#### Network density is higher in 65+ years old

For non-ESBL-EC, resistance associations networks were denser in individuals aged 65 and over than in individuals under 65, with median densities of respectively 0.103 [0.103; 0.105] and 0.088 [0.088; 0.091] in 2022 (Figure 5B). For ESBL-EC, the median network density was not significantly different between 65+ years old, at 0.289 [0.280; 0.301], and –65 years old, at 0.275 [0.261; 0.294]. The ESBL-EC networks of 65+ individuals mainly involved additional associations of cefuroxime with penicillins (amoxicillin, ampicillin), third and fourth generation cephalosporin (cefotaxime, cefepime) and fluoroquinolones (ciprofloxacin, levofloxacin, norfloxacin) (Figure 5A). Similar results were found when analysing data from 2018 to 2021 (Figure S5).

**Figure 5:**
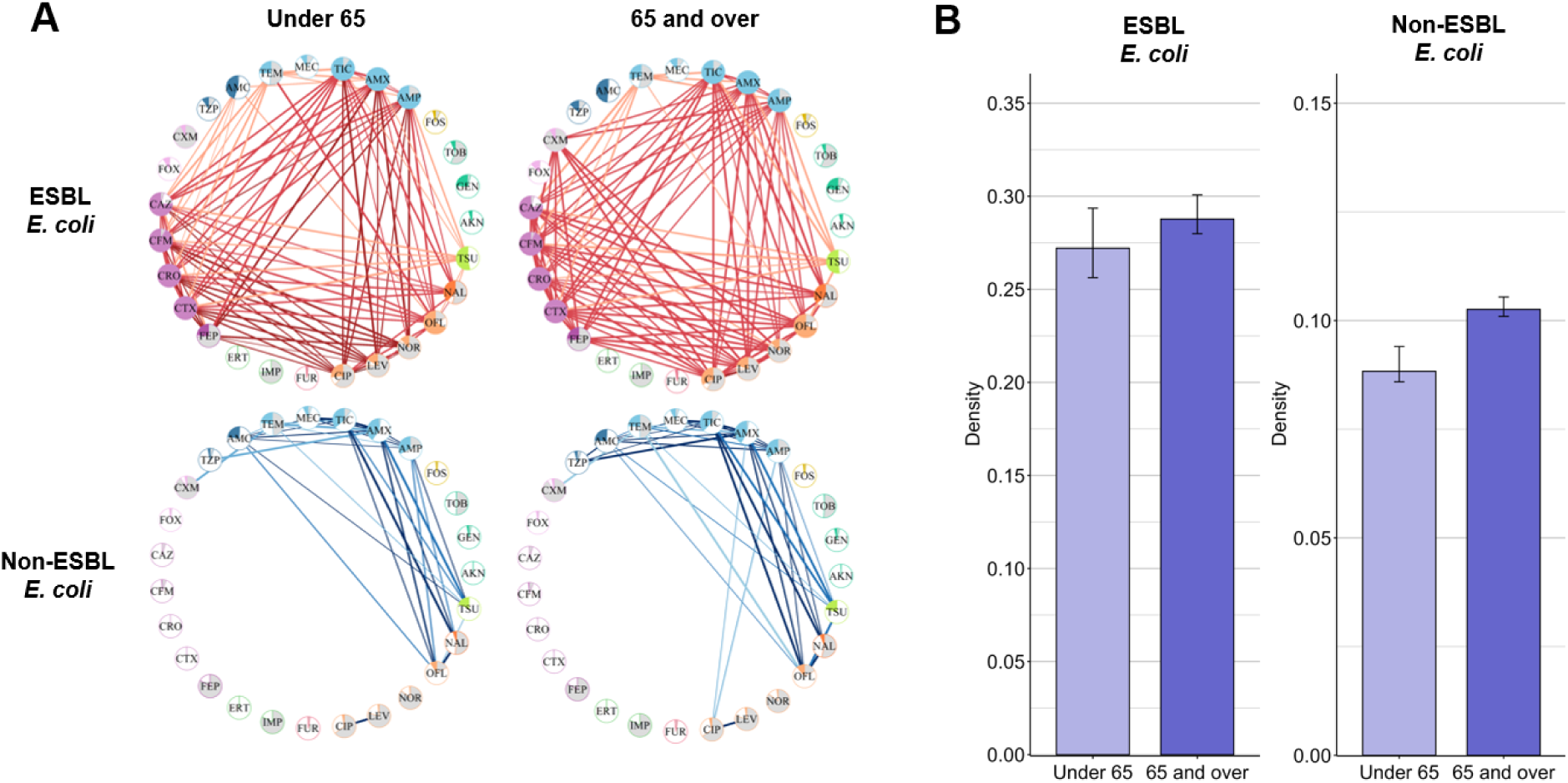
(A) Networks of resistance associations for individuals under 65 and individuals 65 and over in 2022. (B) Barplots of median network density for both age groups. Error bars indicate confidence interval: for individuals over 65, calculated using 10 subsamples randomly generated at the size of the dataset of individuals under 65; for individuals under 65, calculated using 10 bootstrapped datasets from the complete dataset of individuals under 65. See supplementary Table 1 for a complete list of antibiotics with their full name.

#### Network density is higher in men

In 2022 for ESBL-EC, network density was higher in men than in women, with respective medians of 0.305 [0.291; 0.322] and 0.271 [0.266; 0.276] (Figure 6B). The same result was observed in non-ESBL-EC, with median densities of 0.128 [0.125; 0.133] and 0.094 [0.090; 0.096] for men and women respectively. The ESBL-EC networks of men mainly involved additional associations of cefuroxime with penicillins (amoxicillin, ampicillin), third and fourth generation cephalosporin (cefotaxime, cefepime) and fluoroquinolones (ciprofloxacin, levofloxacin, norfloxacin) (Figure 6A). Similar results were found when analysing data from 2018 to 2021 (Figure S6).

**Figure 6:**
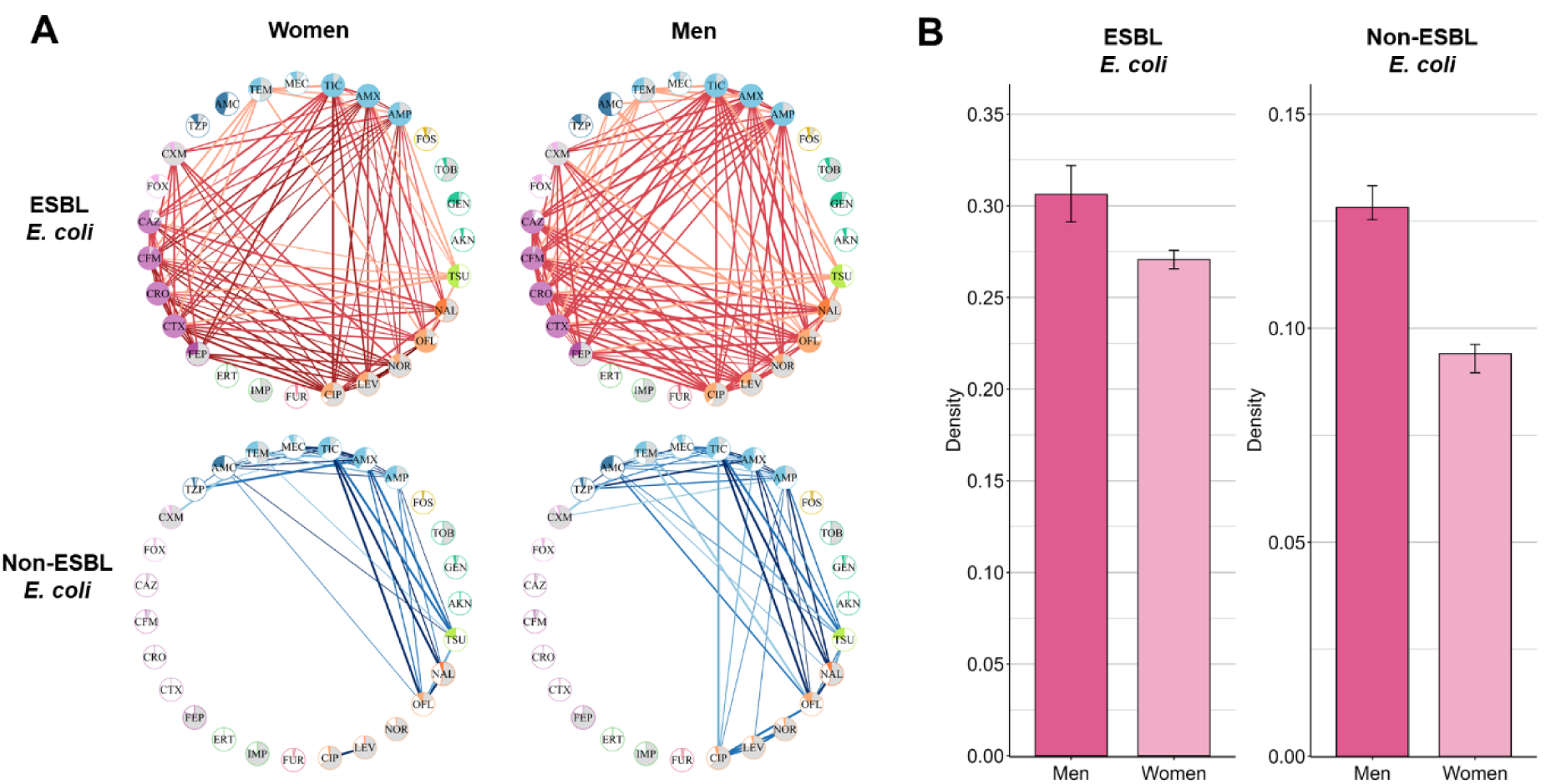
(A) Networks of resistance associations for men and women in 2022. (B) Barplots of the median network density for men and women. Error bars indicate confidence intervals: for women, calculated using 10 datasets generated at the size of the men’s dataset; for men, calculated using 10 bootstrapped datasets from the complete men’s dataset. See supplementary Table 1 for a complete list of antibiotics with their full name.

#### Sensitivity analysis

Figure S7 depicts resistance association networks from 2019 to 2022, obtained with resampled datasets to ensure that the relative contribution of each region is constant over time and proportional to its population. As in the main analysis, networks reconstructed for ESBL-EC isolates were much denser than for non-ESBL-EC. For ESBL-EC, a significant increase in density was again observed over time, from 0.240 to 0.300 (p-value=0.004, Pearson test), although this was not the case any more for non-ESBL-EC.

## Discussion

In this study we identified resistance associations in community-acquired UTI *E. coli* isolates, using a large French surveillance database. Across all years, and as expected, ESBL-EC showed a higher number of resistance associations, particularly involving penicillins, fluoroquinolones and third-generation cephalosporins. In both ESBL and non-ESBL-EC, the number of resistance associations increased from 2018 to 2022, and men and individuals aged 65 and over showed more resistance associations across all years.

We compared the observed distribution of the number of resistances per isolate with a simulated dataset assuming the hypothesis of mutual independence of resistance traits (8) and found significant differences. This finding aligns with Chang et al., 2015, which reported that MDR occurred at frequencies inconsistent with the hypothesis of mutual independence of resistance traits for multiple bacteria including *E. coli* (5). It provides further evidence for biological or genetic mechanisms favouring the co-occurrence of multiple resistance traits in *E. coli*.

Resistance rates were generally low for first-line antibiotics recommended in France for community-acquired UTIs, such as fosfomycin, pivmecillinam or nitrofurantoin (13). Analysis of ESBL-EC resistance associations networks highlighted major associations between penicillins (+/-inhibitors), cephalosporins, fluoroquinolones and cotrimoxazole. This pattern may correspond to CTX-M, the most prevalent ESBL in *E. coli*, as well as TEM or SHV enzymes, which all hydrolyse penicillins and cephalosporins. CTX-M are mostly susceptible to piperacillin-tazobactam (∼80–90%), while this is more variable in TEM/SHV. They are both highly resistant to fluoroquinolones (80–90%), and moderately resistant (∼50%) to cotrimoxazole. Although the mechanisms of resistance to beta-lactams and fluoroquinolones are distinct, fluoroquinolone resistance genes (e.g., qnr or aac(6’)-Ib-cr) are present on mobile genetic elements that also carry beta-lactam resistance genes, which allows multiple resistances to be transmitted jointly (14). Association-set mining may represent an efficient approach to better understand the dynamics of *E. coli* multiresistance patterns and make hypothesis regarding the underlying genetic evolutions, to be confirmed by whole genome sequencing analysis.

Using association-set mining, we found that the number of resistance associations increased from 2018 to 2022 in both ESBL-EC and non-ESBL-EC. Despite this overall increasing trend, a temporary decrease was observed in 2020, consistently with the observed reduction in MDR *E. coli* during the Covid-19 pandemic in France (15). We explored the impact of spatial resistance epidemiology on our findings through a sensitivity analysis using datasets where the number of isolates from each region was proportional to its population. This approach confirmed the nationwide increase of resistance associations within community-acquired ESBL-EC UTIs. However, in this sensitivity analysis, the increase in non-ESBL-EC isolates disappeared. Consequently, the changes in regional relative contributions may have played a role in the temporal increase of the number of resistance associations we observed in non-ESBL-EC isolates. In particular, the contribution of several densely-populated and internationally-connected French regions, including Ile-de-France (Paris area), increased over time. Isolates from these regions may present more resistance associations.

During 2018-2022, the global consumption of antibiotics in primary care decreased in France from 22.9 to 21.6 defined daily doses (DDD) per 10,000 inhabitant per day. This trend was observed for large-spectrum penicillins, cephalosporins and quinolones, but not for associations of penicillins, sulphonamides and trimethoprim which remained stable. In this context, the increasingly dense MDR patterns we observe among community *E. coli* may be driven not only by antibiotic use but also by other multiple human, animal, and environmental factors (16).

Isolates coming from men and individuals older than 65 had more resistance associations, even after adjusting for different sample sizes. This finding is consistent with multiple studies showing that older individuals are particularly exposed to MDR bacteria notably because of immunosenescence, frequent multiple chronic comorbidities and the associated requirement for care. They are more frequently exposed to hospitals and other healthcare facilities, increasing their contact with resistant bacteria (17,18). In contrast, limited data is available regarding gender-specific differences in MDR, although some studies suggested that men have more risks than women to be infected with resistant bacteria (19–21). In our study, men-specific networks had additional associations involving cefuroxime. In hospital settings, cefuroxime is routinely used as surgical prophylaxis to reduce the risk of postoperative infections, particularly in prostate resection, a frequent procedure in older men (22). Moreover, UTIs in men are more commonly associated with structural abnormalities like renal stones and malignancies, which can complicate the treatment, lead to increased antibiotic use and contribute to the development of resistance (23). This could partially explain the additional associations.

Our study is subject to certain limitations that should be taken into account.

First, the size of the PRIMO datasets generally increased every year, notably due to the increase of the scope of the PRIMO mission. However, the 2020 dataset was slightly smaller than the 2019 one (450 135 vs. 450 820 isolates). This could be partially explained by the COVID-19 pandemic which disrupted healthcare systems and microbiological surveillance practices in 2020. Community clinical laboratories prioritized COVID-19 management, potentially reducing their capacity to collect and record AST (24). While network size is known to impact its density (25), in our analyses, we accounted for varying dataset sizes by randomly drawing subsamples at the size of the 2018 dataset.

Second, the PRIMO surveillance system contained isolates that were collected in community clinical laboratories; however, we cannot be certain that it was representative of the total community-acquired *E. coli* UTI in France between 2018 and 2022. Some isolates could have been acquired in healthcare settings, especially for older individuals who are frequently hospitalized. Moreover, while we were unable to collect clinical information, some *E. coli* were potentially isolated from asymptomatic bacteriuria of colonised patients.

Third, our results are valid for *E. coli* isolates only, and not for other Enterobacterales, potentially limiting their generalizability. We chose to focus the study on *E. coli* as it represented 85.0% of all isolates in the dataset.

Fourth, in the PRIMO database, some antibiotics were tested on a very small number of isolates. To minimize bias, we included only antibiotics tested on at least 10% of isolates across all datasets. We found stable results for most antibiotics over time. However, this limitation may explain the changing results regarding ampicillin from 2018 to 2019. In 2018, ampicillin was tested on only 2% of isolates, which likely prevented the algorithm from identifying significant associations involving this antibiotic. By 2019, ampicillin was tested on more isolates, allowing the algorithm to detected such associations. Moreover, the antibiotics tested were determined by the laboratories, and might not align with hospital practices, while consistent with practices in outpatient care.

Fifth, the *Apriori* algorithm is a highly efficient method supported by multiple studies (7–9). However, combining multiple data mining approaches, as recently suggested (9), could have enhanced our understanding of MDR. In addition, *Apriori* generates numerous patterns, requiring careful pruning with appropriate quality measures and cut-off values. While many studies rely solely on support (26), this would have favoured frequent resistance patterns, such as those involving amoxicillin, while overlooking rarer but significant associations. We therefore chose lift as a robust complementary metric (7); however, numerous other metrics or metric combinations could be proposed (27,28), potentially yielding slightly different results. Missing data may also have influenced *Apriori* results by altering resistance prevalence distributions and associations (29), although this was addressed by the use of adapted quality measures (eSup and cLift) (30). Finally, cut-off values also influence pattern selection (26). Our percentile-based approach led us to exclude patterns with a lift below 1, which could have highlighted MDR patterns occurring less frequently than expected.

Despite these limitations, our study provided a novel and detailed analysis of multiresistance patterns in community-acquired *E. coli* UTI collected from a French national surveillance system. We explored the temporal evolution of resistance associations, gender-specific and age-specific differences, which to our knowledge, had not been previously analysed. Our findings confirmed that this method is effective for identifying resistance associations in antibiotic resistance surveillance data. With further research, this work could provide insights for antibiotic stewardship strategies in alignment with known resistance associations in community-acquired *E. coli* UTIs. In the context of rising antibiotic resistance, optimizing the use of current medications is crucial, as few new antibiotics have been developed in the past two decades (31).

Future research could use other machine learning approaches to further analyse the PRIMO datasets and get a deeper understanding of resistance associations. Moreover, it would be interesting to examine the evolution of resistance associations in future years. In addition, extending our approach to other pathogens beyond *E. coli* could offer a broader perspective on multiresistance dynamics. Finally, future work could investigate the potential linkage between these phenotypic resistance associations and genetic co-resistance mechanisms.

## Data Availability

All data produced in the present study are available upon reasonable request to the authors

## List of abbreviations

AMR: Antimicrobial-resistant
AST: Antibiotic susceptibility testing
cLift: conditional lift
DDD: defined daily dose
ESBL-EC: Extended-spectrum beta-lactamase producing *E. coli*
eSup: expected support
MDR: multidrug-resistant
UTI: urinary tract infection

## Declarations

### Availability of data and materials

All code used in the analysis are available in the following GitHub repository: https://github.com/elisehodbert/exploring_MDR_patterns

### Authors’ contributions

Dr Birgand had full access to all of the data in the study and takes responsibility for the integrity of the data and the accuracy of the data analysis.

Concept and design: Hodbert, Lemenand, Temime, Birgand.

Acquisition, analysis, or interpretation of data: Hodbert, Thibaut, Coeffic, Lemenand, Temime, Boutoille, Corvec, Birgand.

Drafting of the manuscript: Hodbert, Birgand, Temime.

Critical revision of the manuscript for important intellectual content: Hodbert, Lemenand, Corvec, Boutoille, Birgand, Temime.

Statistical analysis: Hodbert, Coeffic, Temime Obtained funding: Birgand, Temime.

Administrative, technical, or material support: Birgand, Temime. Supervision: Temime, Lemenand, Birgand.

## French clinical laboratories nationwide network

A. Vrain, LABOUEST, Ancenis ; P. Andorin, BIOLARIS, Laval ; J. Besson, BIOLIANCE, Nantes ; F. Maillet, BIOLOIRE, Nantes ; G. de Gastines, BIORYLIS, La Roche sur Yon ; P.-Y. Léonard, LABORIZON MAINE ANJOU, Le Mans ; M. Guery, SEVRE BIOLOGIE, Les Herbiers ; V. Plong, ACTIV’BIOLAB, Challans ; N. Le Moing, RESEAUBIO, La Chapelle sur Erdre ; F. Kerdavid, ALLIANCE ANABIO, Melesse ; A.-S. Reinhard, BIOCELIANDE, Montauban de Bretagne ; S. Gillard, BIOLOR, Lorient ; B. Guesnon, BIORANCE, Saint Malo ; B. Gestin, LABAZUR, Chateaulin ; H. Banctel, SBL BIO, Saint Brieuc ; D. Laforest, BIOCENTRE, Coutances ; E. Pradier, CARMES, Caen ; S. Arsene, CERBALLIANCE NORMANDIE, Lisieux ; A. Holstein, ABO +, Tours ; D. Bouvet, BIO MEDI QUAL Centre, Vierzon ; B. Dubet, LBM DUBET, Neuville aux bois ; C. Laudignon, MLAB, Saran ; E. Jobert, MIRIALIS, Annecy ; R. Gebeile, DYNABIO, Lyon ; S. Poyet, DYOMEDEA, Lyon ; G. Deleglise, GENBIO, Clermont Ferrand ; N. Lecordier, ANALYSIS 88, Epinal ; S. Fougnot, ATOUTBIO, Nancy ; E. Grandsire, DYNALAB, Romilly sur Seine ; J.-P. Rault, ESPACEBIO, Metz ; G. Defrance, BIOFUTUR, L’isle Adam ; J Cadenet, BIOVSM, Torcy ; L Libier, AX BIO OCEAN, Bayonne ; A Touzalin, BIO17, La Rochelle ; A Allery, BIO86, Poitiers ; H Valade, BIOFFICE, Bordeaux ; G. Payro, CERBALLIANCE-CHARENTE, Saintes ; D. Boraud, EXALAB Groupe LABEXA, Le Haillan ; E. Parisi, VIALLE, Bastia ; F. Alluin, 2A2B, Porto Vecchio ; J. Bayette, LABOSUD, Montpellier ; M.-F. Aran, BIOPOLE66, Perpignan ; P. Stevenin, BIOAXIOME, Avignon ; A. François, BIOESTEREL, Mandelieu-la-Napoule ; G. Gay, LABOSUD PROVENCE, Marseille ; O. Duquesnoy, BIOPATH, Dunkerque ; V. Sainte Rose, BIOLAB MARTINIQUE, Fort de France ; F. Dos Santos, BIOSANTE, Schoelcher. F. Artur, BIOCEANE, Le Havre. E. Mbenga, BIOLAB, Beaune ; A. Desjardins, EVORIAL, Nevers ; M.-C. Paolini, BDCBM5, Besançon ; P. Marchenay, LPA18, Vesoul.

## Group Information

A complete list of the members of the French Clinical Laboratories Nationwide Network appears in Supplement.

## Additional Contributions

Anne Berger-Carbonne, MD, Laetita Gambotti, and Sylvie Maugat, MSc (from Santé Publique France), supported the data. These individuals were not compensated for their contributions.

## References

1. Cosgrove SE. The Relationship between Antimicrobial Resistance and Patient Outcomes: Mortality, Length of Hospital Stay, and Health Care Costs. Clin Infect Dis. 15 janv 2006;42(Supplement_2):S82-9.

2. Holmes AH, Moore LSP, Sundsfjord A, Steinbakk M, Regmi S, Karkey A, et al. Understanding the mechanisms and drivers of antimicrobial resistance. The Lancet. 9 janv 2016;387(10014):176-87.

3. Naghavi M, Vollset SE, Ikuta KS, Swetschinski LR, Gray AP, Wool EE, et al. Global burden of bacterial antimicrobial resistance 1990–2021: a systematic analysis with forecasts to 2050. The Lancet [Internet]. 16 sept 2024 [cité 23 sept 2024];0(0). Disponible sur: https://www.thelancet.com/journals/lancet/article/PIIS0140-6736(24)01867-1/fulltext?dgcid=twitter_organic_gbd24_lancet

4. Murray CJL, Ikuta KS, Sharara F, Swetschinski L, Aguilar GR, Gray A, et al. Global burden of bacterial antimicrobial resistance in 2019: a systematic analysis. The Lancet. 12 févr 2022;399(10325):629-55.

5. Chang HH, Cohen T, Grad YH, Hanage WP, O’Brien TF, Lipsitch M. Origin and proliferation of multiple-drug resistance in bacterial pathogens. Microbiol Mol Biol Rev MMBR. mars 2015;79(1):101-16.

6. Tzouvelekis LS, Markogiannakis A, Psichogiou M, Tassios PT, Daikos GL. Carbapenemases in Klebsiella pneumoniae and other Enterobacteriaceae: an evolving crisis of global dimensions. Clin Microbiol Rev. oct 2012;25(4):682-707.

7. Cazer CL, Al-Mamun MA, Kaniyamattam K, Love WJ, Booth JG, Lanzas C, et al. Shared Multidrug Resistance Patterns in Chicken-Associated Escherichia coli Identified by Association Rule Mining. Front Microbiol [Internet]. 2019 [cité 2 oct 2023];10. Disponible sur: https://www.frontiersin.org/articles/10.3389/fmicb.2019.00687

8. Cazer CL, Westblade LF, Simon MS, Magleby R, Castanheira M, Booth JG, et al. Analysis of Multidrug Resistance in Staphylococcus aureus with a Machine Learning-Generated Antibiogram. Antimicrob Agents Chemother. 18 mars 2021;65(4):10.1128/aac.02132-20.

9. Mosaddegh A, Angel CC, Craig M, Cummings KJ, Cazer CL. An exploration of descriptive machine learning approaches for antimicrobial resistance: Multidrug resistance patterns in Salmonella enterica. Prev Vet Med. sept 2024;230:106261.

10. Lemenand O, Thibaut-Jovelin S, Coeffic T, Caillon J. PRIMO : Résultats 2022 - Surveillance de la résistance bactérienne aux antibiotiques en soins de ville et en Ehpad [Internet]. Repias : Réseau de Prévention des Infections Associées aux Soins. 2024 [cité 20 déc 2024]. Disponible sur: https://www.preventioninfection.fr/actualites/primo-resultats-2022-surveillance-de-la-resistance-bacterienne-aux-antibiotiques-en-soins-de-ville-et-en-ehpad/

11. Cattoir V. Comité de l’Antibiograme de la Société Française de Microbiologie [Internet]. Société Française de Microbiologie. 2022 [cité 5 févr 2025]. Disponible sur: https://www.sfm-microbiologie.org/boutique/_comite-de-lantibiogramme-de-la-sfm-ca-sfm-v1-0-juin-2024/

12. Dongre J, Prajapati GL, Tokekar SV. The role of Apriori algorithm for finding the association rules in Data mining. In: 2014 International Conference on Issues and Challenges in Intelligent Computing Techniques (ICICT) [Internet]. Ghaziabad, India: IEEE; 2014 [cité 4 juin 2024]. p. 657-60. Disponible sur: http://ieeexplore.ieee.org/document/6781357/

13. Caron F, Galperine T, Flateau C, Azria R, Bonacorsi S, Bruyère F, et al. Practice guidelines for the management of adult community-acquired urinary tract infections. Med Mal Infect. 2018;48(5):327-58.

14. Strahilevitz J, Jacoby GA, Hooper DC, Robicsek A. Plasmid-Mediated Quinolone Resistance: a Multifaceted Threat. Clin Microbiol Rev. oct 2009;22(4):664-89.

15. Lemenand O, Coeffic T, Thibaut S, Cotinat MC, Caillon J, Birgand G. Decreasing proportion of extended-spectrum beta-lactamase among E. coli infections during the COVID-19 pandemic in France. J Infect. 1 déc 2021;83(6):664-70.

16. Paumier A, Asquier-Khati A, Thibaut S, Coeffic T, Lemenand O, Larramendy S, et al. Assessment of Factors Associated With Community-Acquired Extended-Spectrum β-Lactamase-Producing Escherichia coli Urinary Tract Infections in France. JAMA Netw Open. 1 sept 2022;5(9):e2232679.

17. Gajdács M, Ábrók M, Lázár A, Burián K. Urinary Tract Infections in Elderly Patients: A 10-Year Study on Their Epidemiology and Antibiotic Resistance Based on the WHO Access, Watch, Reserve (AWaRe) Classification. Antibiotics. sept 2021;10(9):1098.

18. Theodorakis N, Feretzakis G, Hitas C, Kreouzi M, Kalantzi S, Spyridaki A, et al. Antibiotic Resistance in the Elderly: Mechanisms, Risk Factors, and Solutions. Microorganisms. oct 2024;12(10):1978.

19. Brandl M, Hoffmann A, Willrich N, Reuss A, Reichert F, Walter J, et al. Bugs That Can Resist Antibiotics but Not Men: Gender-Specific Differences in Notified Infections and Colonisations in Germany, 2010–2019. Microorganisms [Internet]. 2021 [cité 18 nov 2024];9. Disponible sur: https://consensus.app/papers/bugs-that-resist-antibiotics-genderspecific-differences-brandl/2b9390f598b05d908488d513c5c4a2b3/

20. Khanal N, Cortie CH, Story C, Jones S, Mansfield KJ, Miyakis S, et al. Multidrug resistance in urinary E. coli higher in males compared to females. BMC Urol. 18 nov 2024;24(1):255.

21. Costa T, Linhares I, Ferreira R, Neves J, Almeida A. Frequency and Antibiotic Resistance of Bacteria Implicated in Community Urinary Tract Infections in North Aveiro Between 2011 and 2014. Microb Drug Resist. mai 2018;24(4):493-504.

22. Ferrie B, Scott R. Prophylactic cefuroxime in transurethral resection. Urol Res. 2004;12:279-81.

23. Wagenlehner FME, Weidner W, Pilatz A, Naber KG. Urinary tract infections and bacterial prostatitis in men. Curr Opin Infect Dis. févr 2014;27(1):97.

24. Durant TJS, Peaper DR, Ferguson D, Schulz WL. Impact of COVID-19 Pandemic on Laboratory Utilization. J Appl Lab Med. 1 nov 2020;5(6):1194-205.

25. Snijders T, Borgatti S. Non-Parametric Standard Errors and Tests for Network Statistics. Connections. janv 1999;22.

26. Martínez-Ballesteros M, Martínez-Álvarez F, Lora AT, Santos JCR. Selecting the best measures to discover quantitative association rules. Neurocomputing. 2014;126:3-14.

27. Datta S, Mali K. Significant Association Rule Mining with High Associability. In: 2021 5th International Conference on Intelligent Computing and Control Systems (ICICCS) [Internet]. 2021 [cité 12 déc 2024]. p. 1159-64. Disponible sur: https://ieeexplore.ieee.org/document/9432237

28. Tan PN, Kumar V, Srivastava J. Selecting the right interestingness measure for association patterns. In: Proceedings of the eighth ACM SIGKDD international conference on Knowledge discovery and data mining [Internet]. New York, NY, USA: Association for Computing Machinery; 2002 [cité 28 mars 2024]. p. 32-41. (KDD ‘02). Disponible sur: 10.1145/775047.775053

29. Brown ML, Kros JF. Data mining and the impact of missing data. Ind Manag Data Syst. 1 janv 2003;103(8):611-21.

30. Calders T, Goethals B, Mampaey M. Mining itemsets in the presence of missing values: 22nd ACM Symposium on Applied Computing (SAC 2007). Cho Y, Wainwright RL, Haddad H, Shin SY, Koo YW, éditeurs. Proc 22nd Annu ACM Symp Appl Comput SAC 2007 11-15 March 2007 Seoul Korea. 2007;404-8.

31. Renwick MJ, Brogan DM, Mossialos E. A systematic review and critical assessment of incentive strategies for discovery and development of novel antibiotics. J Antibiot (Tokyo). févr 2016;69(2):73-88.

