## Supplementary material for "Exploring multidrug resistance patterns in community-acquired *E. coli* urinary tract infections with machine learning"

### Table of contents

**Supplementary table 1.** Selection of antibiotics for the analyses

**Supplementary text 1.** Basic concepts for identifying frequent resistance patterns.

**Supplementary text 2.** Creation of datasets with a proportional number of isolates

**Figure S1.** Spatial distribution of the samples

**Figure S2.** Individual antibiotic resistance prevalence from 2018 to 2021

**Figure S3.** Distribution of the number of resistances per isolate in the observed and simulated datasets in all isolates for 2018, 2019, 2020 and 2021.

**Figure S4.** Distribution of the number of resistances per isolate in the observed and simulated datasets in ESBL and non-ESBL E. coli for (A) 2018, (B) 2019, (C) 2020, 2021 (D) and 2022 (E).

**Figure S5.** Results of analyses conducted with isolates from individuals under 65 and individuals 65 and over, from 2018 to 2021.

**Figure S6.** Results of analyses conducted with isolates from men and women, from 2018 to 2021.

**Figure S7.** Results of sensitivity analyses accounting for the varying contribution of each region from 2019 to 2021

### Supplementary table 1. Selection of antibiotics for the analyses

| **Class** | **Antibiotic** | **Abv.** | **Mean testing rate across all years** | **Selected?** | **If not selected, reason** |
| --- | --- | --- | --- | --- | --- |
| Aminoglycoside | Amikacin | AKN | 99.1% | yes |  |
|  | Gentamicin | GEN | 93.0% | yes |  |
|  | Tobramycin | TOB | 67.4% | yes |  |
| Other | Fosfomycin | FOS | 97.3% | yes |  |
| Large spectrum penicillin | Ampicillin | AMP | 56.2% | yes |  |
|  | Amoxicillin | AMX | 95.9% | yes |  |
|  | Mecillinam | MEC | 95.1% | yes |  |
|  | Temocillin | TEM | 49.6% | yes |  |
|  | Ticarcillin | TIC | 85.7% | yes |  |
|  | Piperacillin | PIP | 13.2% | no | Antibiotic rarely used in routine outpatient care |
| Penicillin + beta-lactamase inhibitor | Amoxicillin + Clavulanic acid | AMC | 97.5% | yes |  |
|  | Piperacillin +Tazobactam | TZP | 91.9% | yes |  |
|  | Ticarcillin+clavulanic acid | TCC | 7.8% | no | <10% isolates tested across all years |
| First generation cephalosporin | Cefalexin | CN | 5.7% | no | <10% isolates tested across all years |
| Second generation cephalosporin | Cefuroxime | CXM | 12.9% | yes |  |
|  | Cefoxitin | FOX | 98.1% | yes |  |
| Third generation cephalosporin | Ceftazidime | CAZ | 92.8% | yes |  |
|  | Cefixime | CFM | 91.7% | yes |  |
|  | Ceftriaxone | CRO | 96.8% | yes |  |
|  | Cefotaxime | CTX | 86.0% | yes |  |
| Forth generation cephalosporin | Cefepime | FEP | 19.9% | yes |  |
| Carbapenem | Ertapenem | ERT | 98.2% | yes |  |
|  | Imipenem | IMP | 20/6% | yes |  |
|  | Meropenem | MEM | 11.3% | no | Antibiotic rarely used in routine outpatient care |
| Nitrofuran derivative | Nitrofurantoin | FUR | 97.3% | yes |  |
| Fluoroquinolone | Ciprofloxacin | CIP | 44.6% | yes |  |
|  | Levofloxacin | LEV | 17.2% | yes |  |
|  | Norfloxacin | NOR | 24.0% | yes |  |
|  | Ofloxacin | OFL | 71.5% | yes |  |
| Other quinolone | Nalidixic acid | NAL | 74.3% | yes |  |
| Trimethoprim/Sulfamethoxazole | Trimethoprim / Sulfamethoxazole | TSU | 98.6% | yes |  |
| Monobactams | Aztreonam | AZT | 10.5% | no | Antibiotic rarely used in routine outpatient care |
| Polymixins | Colistin | CS | 2.6% | no | <10% isolates tested across all years |
| Glycylcycline | Tigecycline | TGC | 7.7% | no | <10% isolates tested across all years |
| Diaminopyrimidines | Trimethoprim | TMP | 11.9% | no | Antibiotic rarely used in routine outpatient care |

### Supplementary text 1. Identifying and selecting resistance patterns in an antibiotic resistance surveillance dataset.

#### Basic concepts for identifying frequent resistance patterns

Association-set mining is an unsupervised machine learning technique that can identify patterns of associations between elements in a large dataset. We use this method to identify patterns of multiresistance, which are defined as combinations of two or more resistance traits. Given that our binary datasets include 27 distinct antibiotics, there are 2^27^-1 distinct possible resistance association sets. Hence, exploring these sets and their characteristics may be very computer-intensive. Several machine learning algorithms have been developed in this regard. Here, we chose *Apriori*, which is the most commonly used due to its simplicity and robustness in handling large datasets (12).

*Apriori* takes as input a dataset containing binary variables (presence/absence). The algorithm efficiently explores the dataset to identify patterns that meet or exceed a defined frequency called “minimal support”, based on the principle that every subset of a frequent pattern is inherently frequent. The process starts by examining simple patterns of length 1 (individual items), keeping only those whose support exceeds a specified threshold. The algorithm then considers more complex patterns that include the simpler, frequent ones. The algorithm outputs the frequent patterns or associations among the elements. For our analysis, we set the initial minimum support threshold at 0.01 for ESBL *E. coli* and 0.001 for non-ESBL *E. coli*. These thresholds were chosen to balance the need to capture as many potentially important patterns as possible with our technical limitations.

The algorithm usually identifies a large number of patterns, depending on the size of data and the prevalence of antibiotic resistance. This results in a substantial number of “false positive” patterns, which do not reflect true associations between resistance. Therefore, in order to spot the most interesting patterns, we describe and prune them using appropriate metrics.

#### Describing the resistance patterns using measures of interest

Several metrics have been proposed to describe patterns, each with varying relevance depending on the domain of application (28). In this study, we used support and lift. Support measures the pattern frequency within the dataset. It is calculated as the proportion of isolates that are resistant to all antibiotics of that pattern among all isolates in the dataset. Lift assesses the frequency of a pattern relative to its expected frequency under the assumption that resistance to antibiotics within the pattern are independent. It is calculated by dividing the pattern’s support by the expected support under the assumption of mutual independence of resistance traits in the pattern, which is the product of the individual supports of each antibiotic in the pattern.

Our dataset contains missing data with varying proportions depending on antibiotics, due to very different antibiotic testing rates. Therefore we used adapted versions of support and lift, called respectively expected support (eSup) and conditional lift (cLift), that account for missing data (8). eSup is calculated as the proportion of isolates that are resistant to all antibiotics in the pattern, divided by the proportion of isolates for which AST data is available for all antibiotics in the pattern. eSup ranges from 0 to 1, where higher values correspond to more frequent patterns.

$$eSup\left( X \right)=\frac{Support(X)}{Support\left( X \right)+Support(\neg X)}$$

Where X is a pattern of resistance and ¬𝑋 represents all patterns other than X. Support(X) counts isolates that contain pattern X, and Support(¬𝑋) counts isolates that definitely do not contain pattern X.

cLift is calculated as the support of the resistance pattern conditional on all antibiotics in the pattern being tested, meaning no missing susceptibility results, divided by the support that would be expected if all resistance traits in the pattern were mutually independent. A cLift value greater than 1 indicates that the individual resistances within the pattern are positively associated (that is, the pattern occurs more frequently than predicted by chance if the resistances were independent), while a value less than 1 indicates a negative association.

$$cLift\left( X | X* \right)= \frac{Support(X|X*)}{\prod_{x\in X} Support(x)}$$

Where X is a pattern of resistance composed of individual resistances x, and X* identifies isolates for which there is no missing testing data for all resistances included in pattern X.

#### Pruning resistance patterns

We pruned patterns by calculating cut-off values based on the method of (8). We applied the *Apriori* algorithm to the datasets previously simulated under H0. Patterns identified in these datasets are inherently random. For each year and phenotype dataset, we calculated the value of eSup and cLift below which 95% of these random patterns fell, and used these values as cut-offs. In the observed datasets, we pruned patterns that had either eSup or cLift values below the respective cut-off values.

### Supplementary text 2. Creation of regionally proportional datasets

To account for the varying contributions of each French region, we created datasets with isolate counts proportional to the population of each region for each year and phenotype subset. In 2018, only 530 isolates were collected from Ile-de-France, which we deemed insufficient for robust regional representation. Therefore, we focused on data from 2019 to 2022.

We collected regional population data for metropolitan France from INSEE (<https://www.insee.fr/fr/statistiques/6013844?sommaire=6011075>). For each region, we calculated the ratio of isolates to regional population. Below is the data for 2019:

| Region | Number of isolates | Regional population | $\frac{Number of isolates}{Population of the region}$ |
| --- | --- | --- | --- |
| Auvergne-Rhône-Alpes | 37 730 | 8 042 936 | 4,69E-03 |
| Bourgogne-Franche-Comté | 31 628 | 2 805 580 | 1,13E-02 |
| Bretagne | 41 509 | 3 354 854 | 1,24E-02 |
| Centre-Val de Loire | 15 465 | 2 573 180 | 6,01E-03 |
| Corse | 43 55 | 340 440 | 1,28E-02 |
| Grand Est | 37 455 | 5 556 219 | 6,74E-03 |
| Hauts-de-France | 24 662 | 6 004 947 | 4,11E-03 |
| Île-de-France | 18 481 | 12 262 544 | 1,51E-03 |
| Normandie | 31 754 | 3 325 032 | 9,55E-03 |
| Nouvelle-Aquitaine | 58 994 | 6 010 289 | 9,82E-03 |
| Occitanie | 43 443 | 5 933 185 | 7,32E-03 |
| Pays de la Loire | 59 810 | 3 806 461 | 1,57E-02 |
| Provence-Alpes-Côte d'Azur | 45 559 | 5 081 101 | 8,97E-03 |

The Île-de-France region displayed the lowest isolate-to-population ratio, with 18,481 isolates for 12,262,544 residents. We included all Île-de-France isolates were included in our sampled dataset, and we used a cross product to calculate the number of isolates in the sampled dataset, with the following formula:

$$Isolates in sampled dataset= \frac{Isolates in Ile-de-France*Total population of France}{Population of Ile-de-France}$$

This gave 98 108 isolates in the sampled dataset. We then calculated the isolate count for each region using the proportional formula:

$$Isolates from region= \frac{Isolates in sampled dataset*Population of region}{Total population of France}$$

The resulting number of isolates for each region is summarized below:

| Region | Isolates in sampled dataset |
| --- | --- |
| Auvergne-Rhône-Alpes | 12 122 |
| Bourgogne-Franche-Comté | 4 228 |
| Bretagne | 5 056 |
| Centre-Val de Loire | 3 878 |
| Corse | 513 |
| Grand Est | 8 374 |
| Hauts-de-France | 9 050 |
| Île-de-France | 18 481 |
| Normandie | 5 011 |
| Nouvelle-Aquitaine | 9 058 |
| Occitanie | 8 942 |
| Pays de la Loire | 5 737 |
| Provence-Alpes-Côte d'Azur | 7 658 |

For each year from 2019 to 2022, we randomly sampled the specified number of isolates for each region from the original dataset; and then concatenated them into a single dataset per year/phenotype combination. To minimize potential biases related to dataset size, we created datasets of the same size from 2019 to 2022. To account for variability due to random sampling, we repeated this process 10 times. These datasets were then analyzed by association-set mining.

### Figure S1. Spatial distribution of the samples

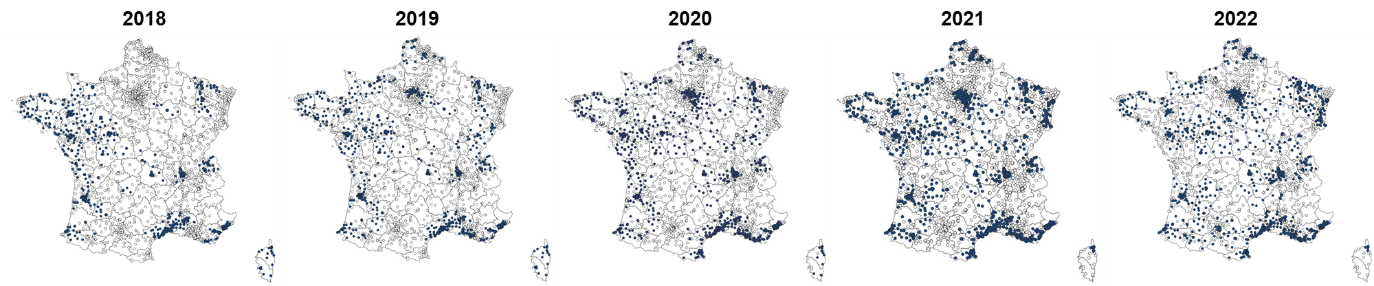

A. Laboratories participating in the PRIMO Mission from 2018 to 2022. Blue dots and white dots represent respectively participating and non-participating laboratories.

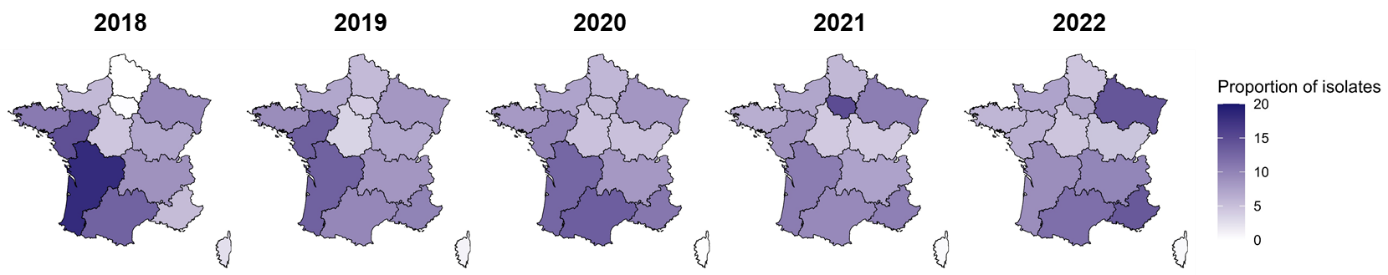

B. Proportion of isolates per administrative region from 2018 to 2022

### Figure S2. Individual antibiotic resistance prevalence in (A) 2018, (B) 2019, (C) 2020 and (D) 2021.

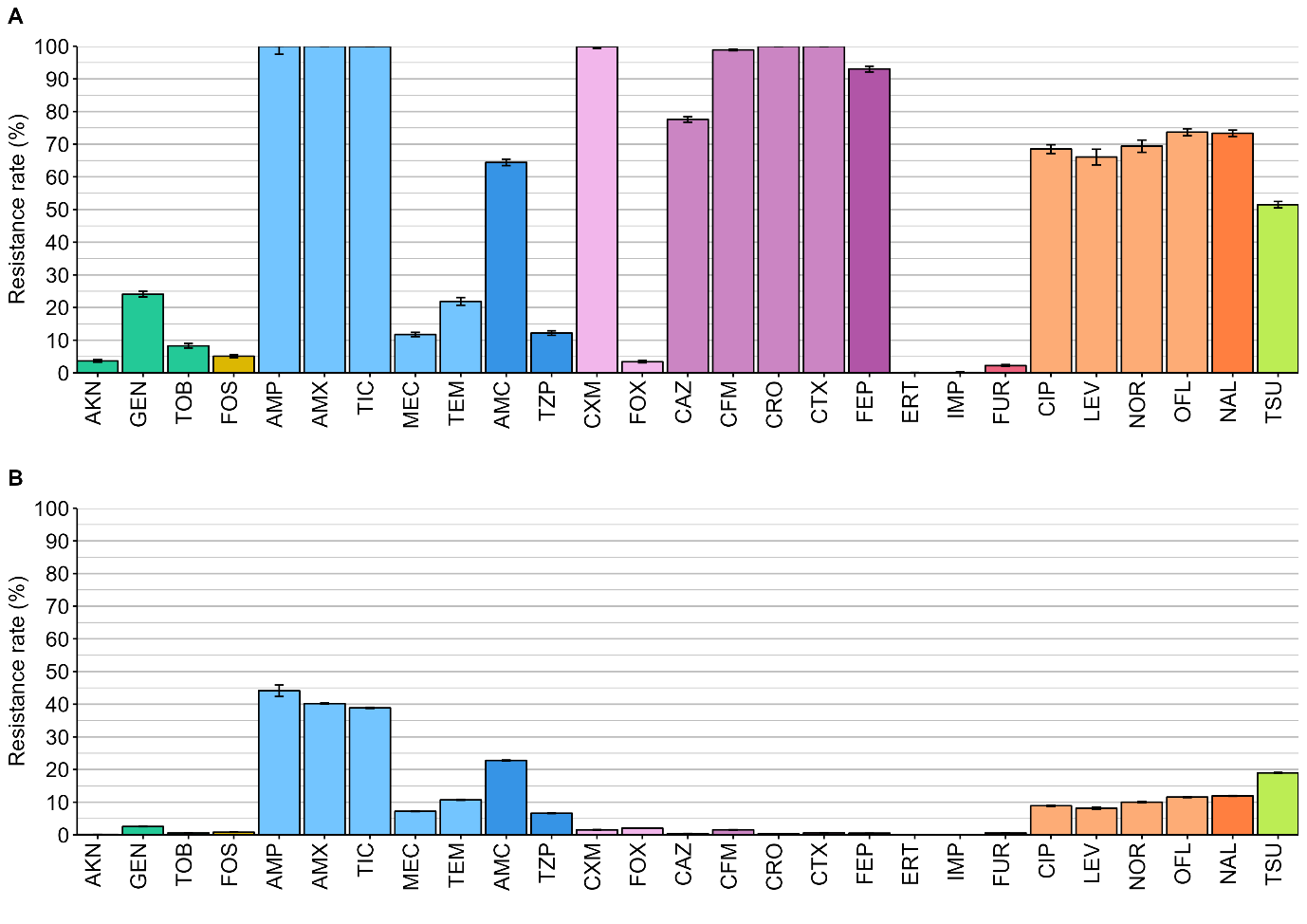

A. Individual antibiotic resistance prevalence in 2018 for ESBL (A) and non-ESBL (B) isolates

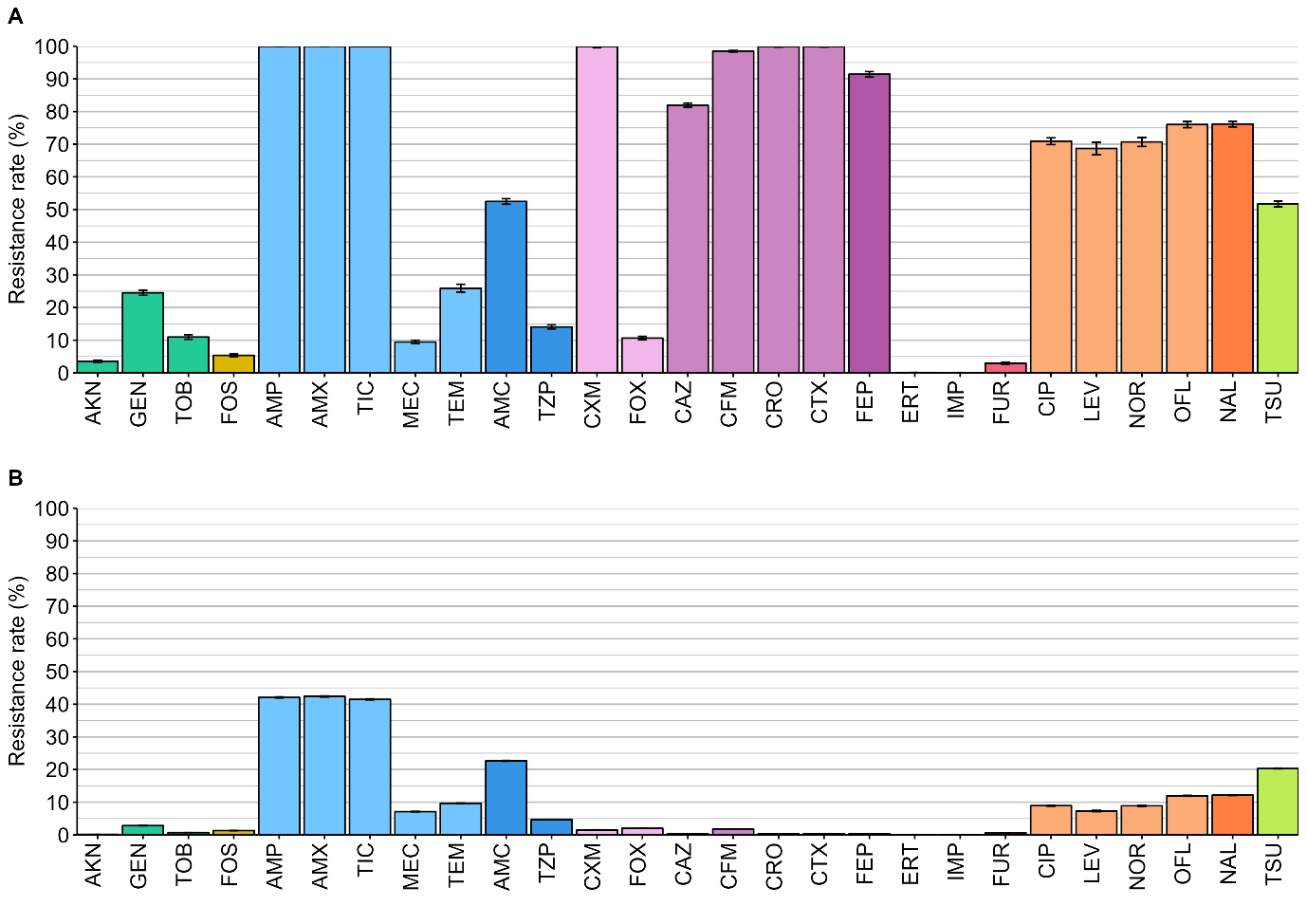

B. Individual antibiotic resistance prevalence in 2019 for ESBL (A) and non-ESBL (B) isolates

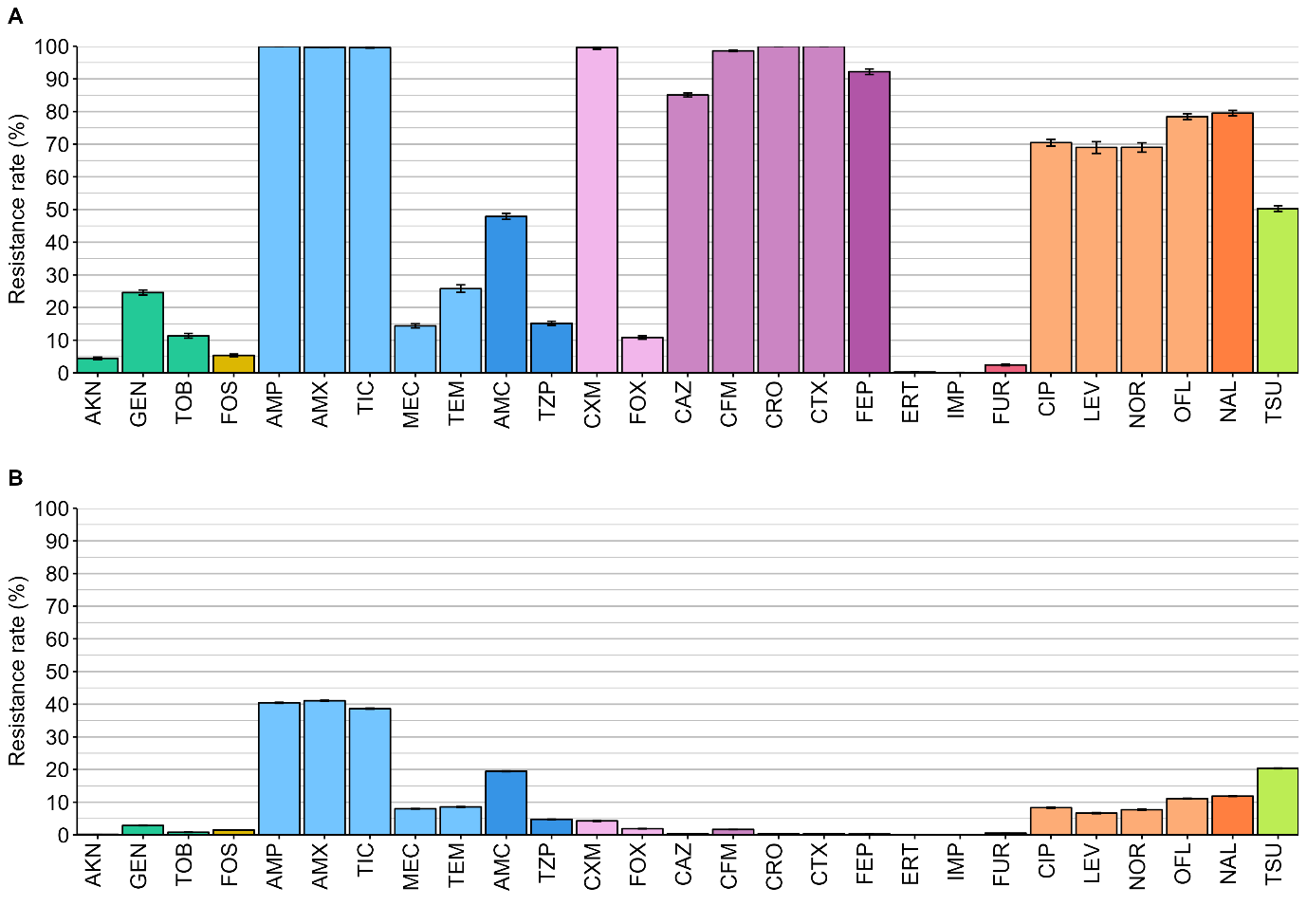

C. Individual antibiotic resistance prevalence in 2020 for ESBL (A) and non-ESBL (B) isolates

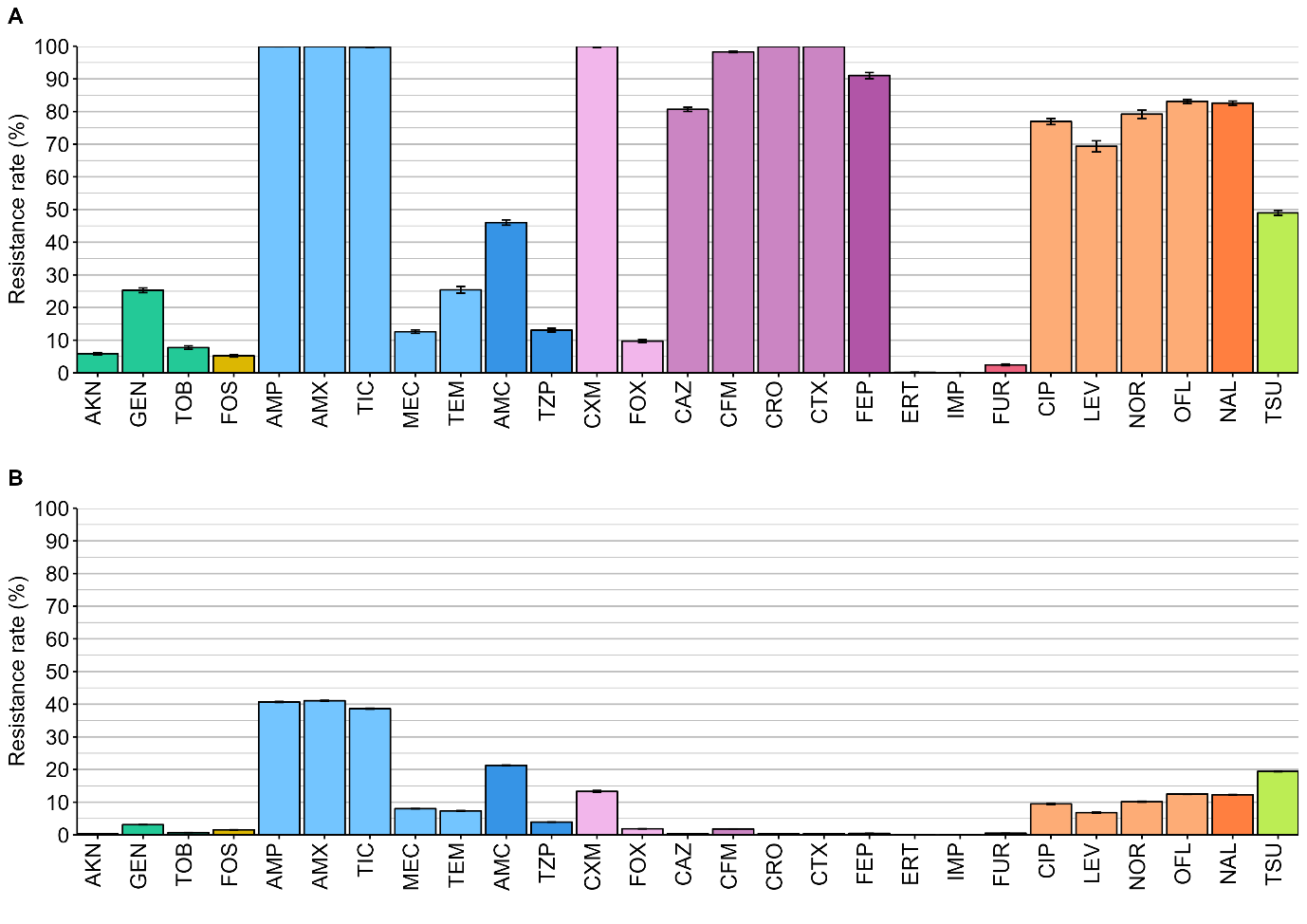

D. Individual antibiotic resistance prevalence in 2021 for ESBL (A) and non-ESBL (B) isolates

### Figure S3. Distribution of the number of resistances per isolate in the observed and simulated datasets in all isolates for (A) 2018, (B) 2019, (C) 2020 and (D) 2021.

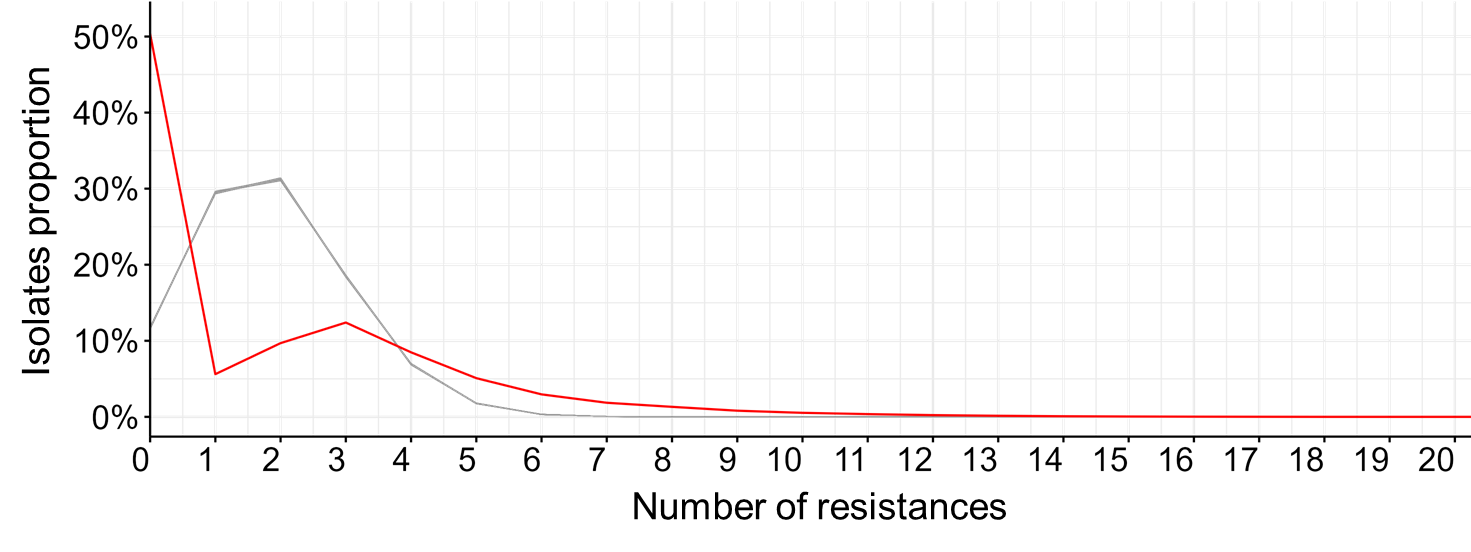

A. Distribution of the number of resistances per isolate in the observed and simulated datasets in 2018 in all isolates. (p-value = 0.02, Kolmogorov-Smirnov test)

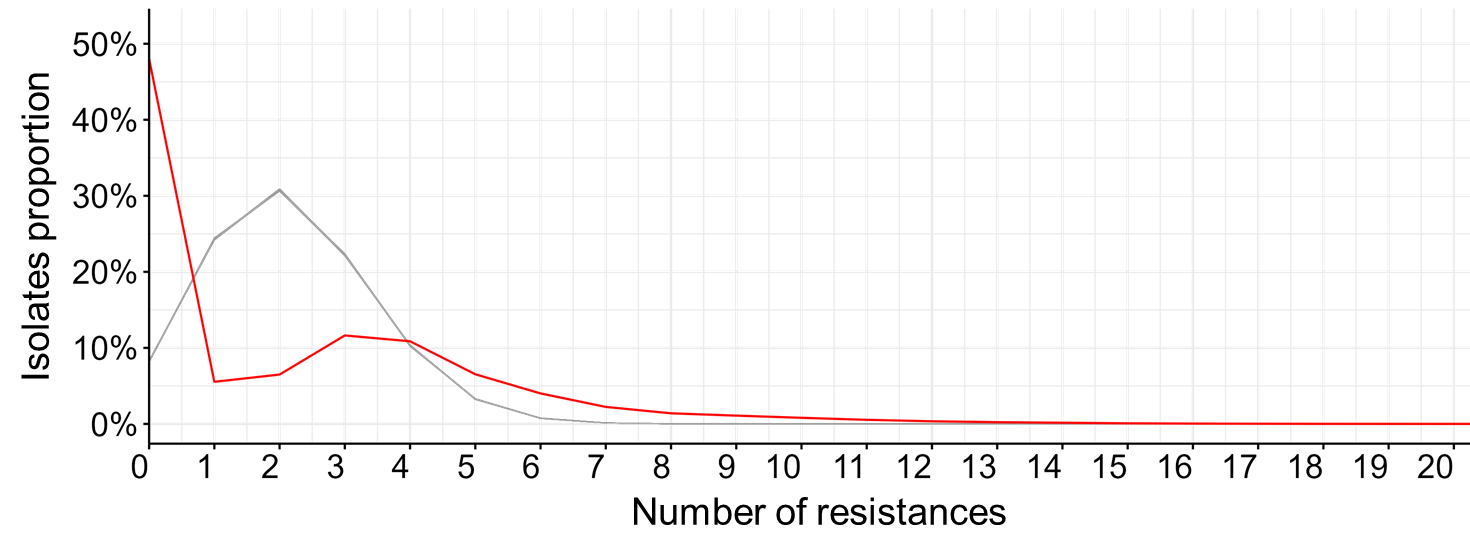

B. Distribution of the number of resistances per isolate in the observed and simulated datasets in 2019 in all isolates. (p-value = 0.02, Kolmogorov-Smirnov test)

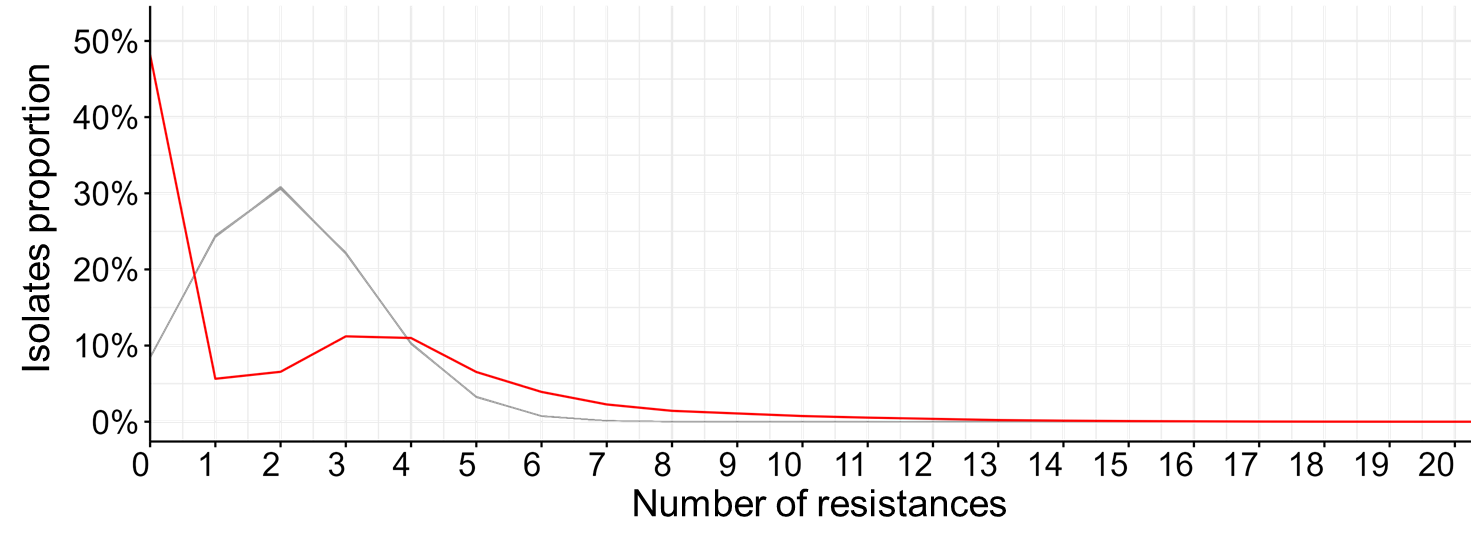

C. Distribution of the number of resistances per isolate in the observed and simulated datasets in 2020 in all isolates. (p-value = 0.01, Kolmogorov-Smirnov test)

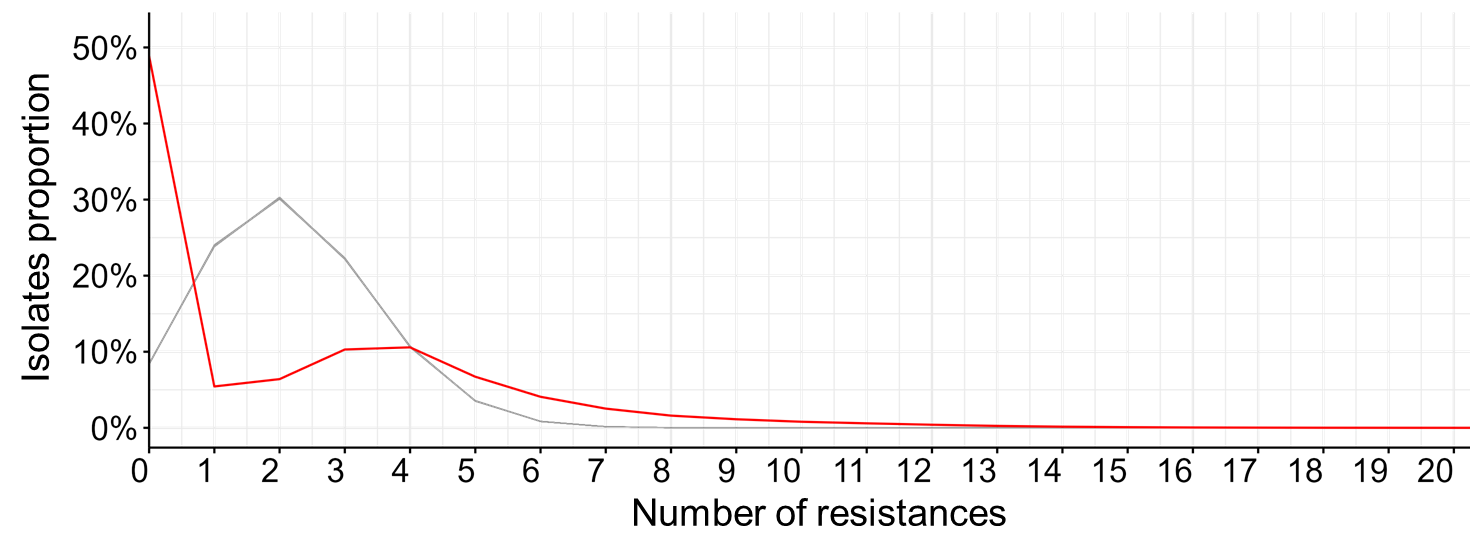

D. Distribution of the number of resistances per isolate in the observed and simulated datasets in 2021 in all isolates. (p-value = 7,89.10^-4^, Kolmogorov-Smirnov test)

### Figure S4. Distribution of the number of resistances per isolate in the observed and simulated datasets in ESBL and non-ESBL *E. coli* for (A) 2018, (B) 2019, (C) 2020, 2021 (D) and 2022 (E).

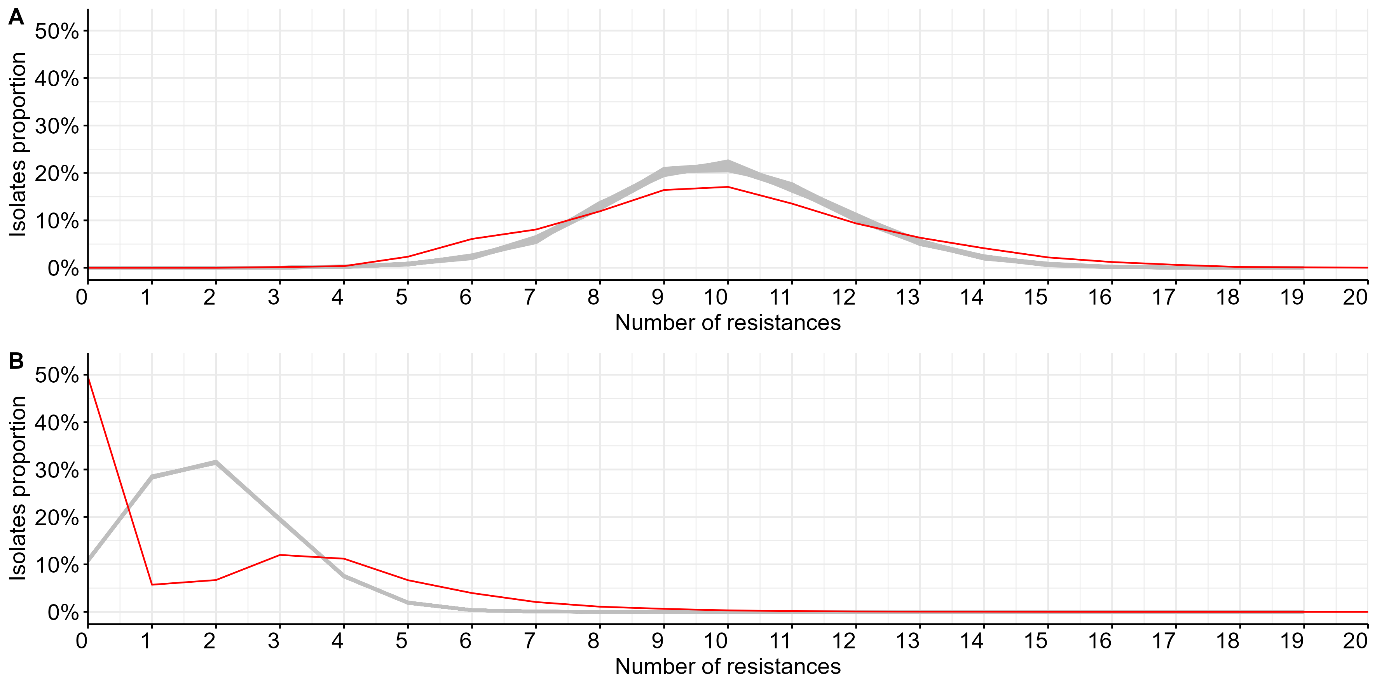

A. Distribution of the number of resistances per isolate in the observed and simulated datasets in 2018 in ESBL (A) and non-ESBL (B) isolates. (ESBL-EC : p-value = 0.67 ; non-ESBL-EC : p-value = 6.00.10^-3^)

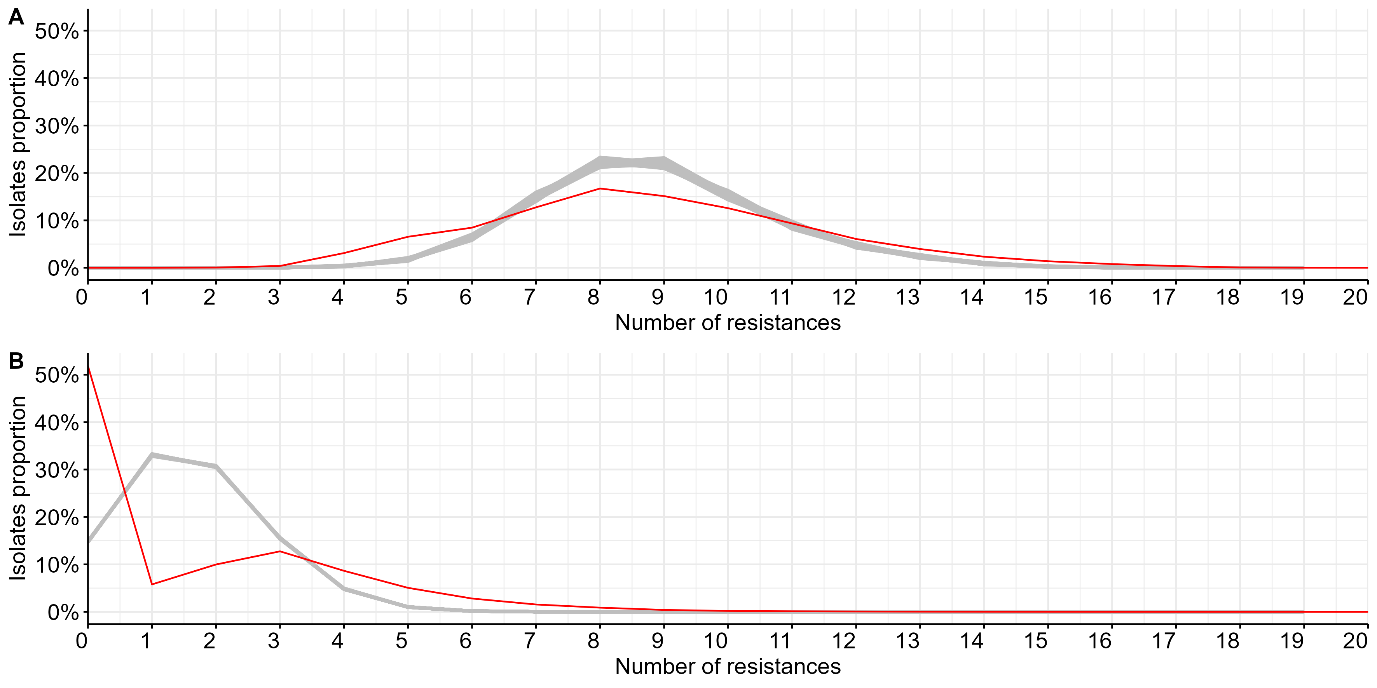

B. Distribution of the number of resistances per isolate in the observed and simulated datasets in 2019 in ESBL (A) and non-ESBL (B) isolates. (ESBL-EC : p-value = 0.88 ; non-ESBL-EC : p-value = 0.03)

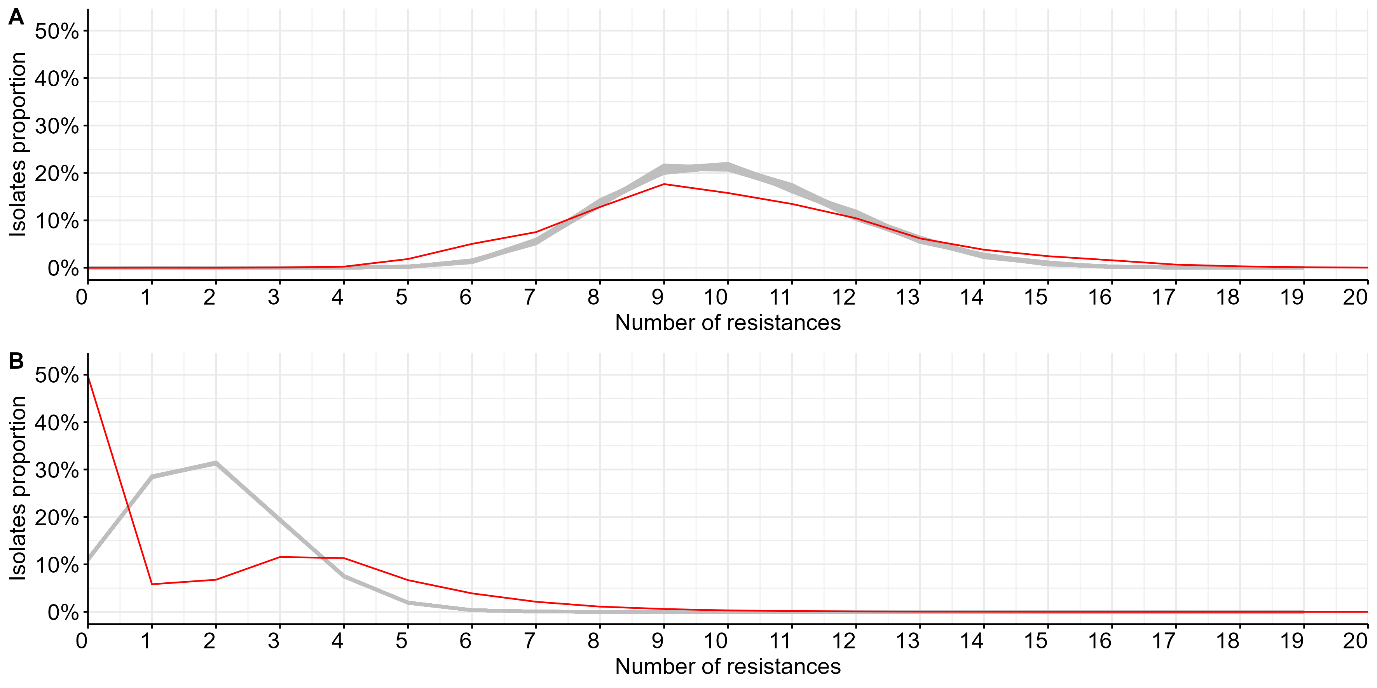

C. Distribution of the number of resistances per isolate in the observed and simulated datasets in 2020 in ESBL (A) and non-ESBL (B) isolates. (ESBL-EC : p-value = 0.47 ; non-ESBL-EC : p-value = 0.02)

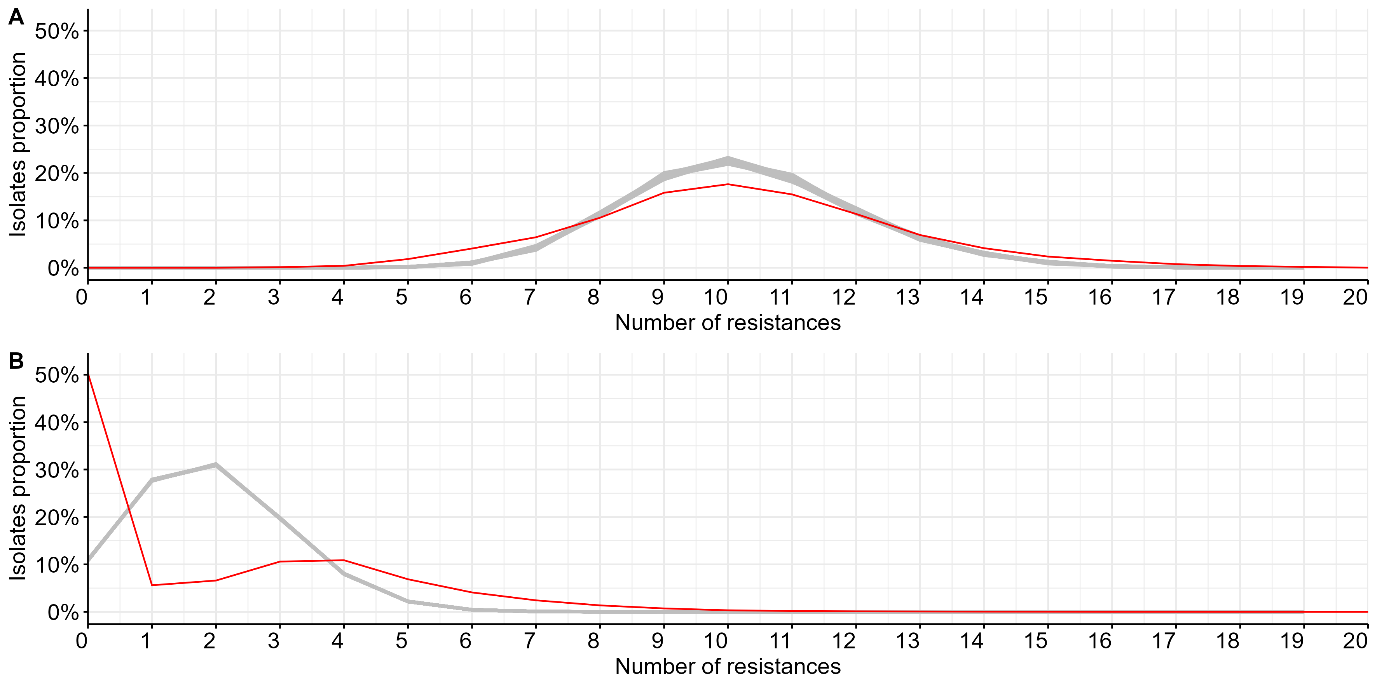

D. Distribution of the number of resistances per isolate in the observed and simulated datasets in 2021 in ESBL (A) and non-ESBL (B) isolates. (ESBL-EC : p-value = 0.48 ; non-ESBL-EC : p-value = 0.001)

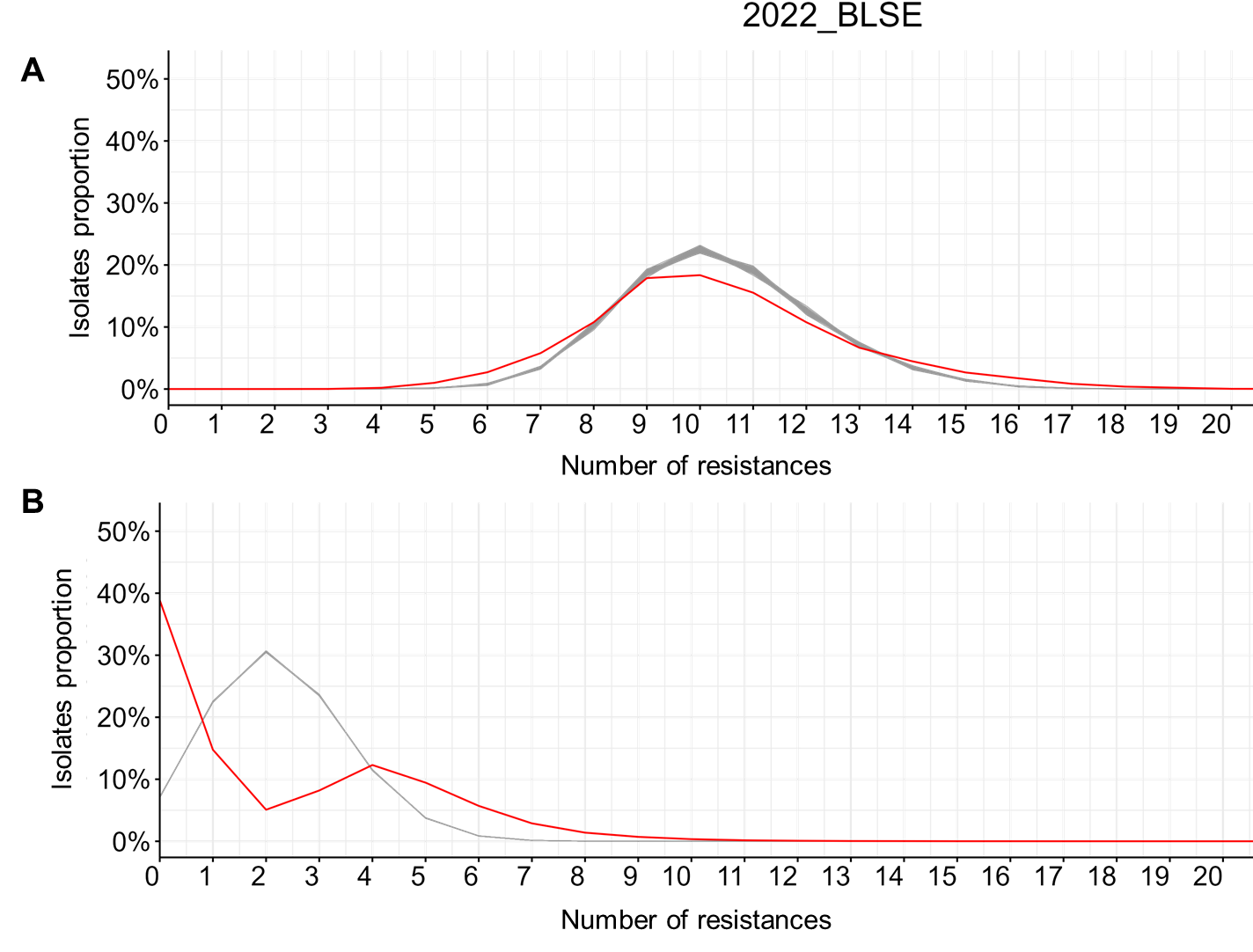

E. Distribution of the number of resistances per isolate in the observed and simulated datasets in 2022 in ESBL (A) and non-ESBL (B) isolates. (ESBL-EC : p-value = 0.70 ; non-ESBL-EC : p-value=0.02)

### Figure S5. Results of analyses conducted with isolates from individuals (A) under 65 and (B) 65 and over, from 2018 to 2021.

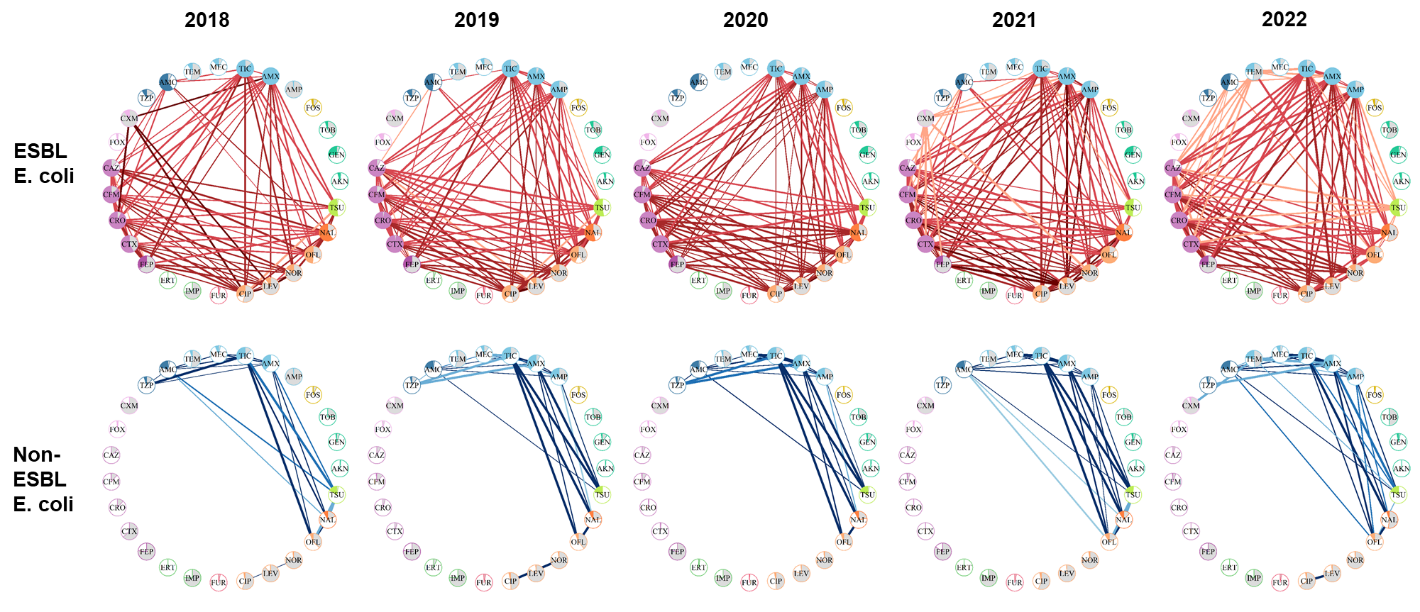

A. Results of analyses conducted with isolates coming from individuals under 65 from 2018 to 2021

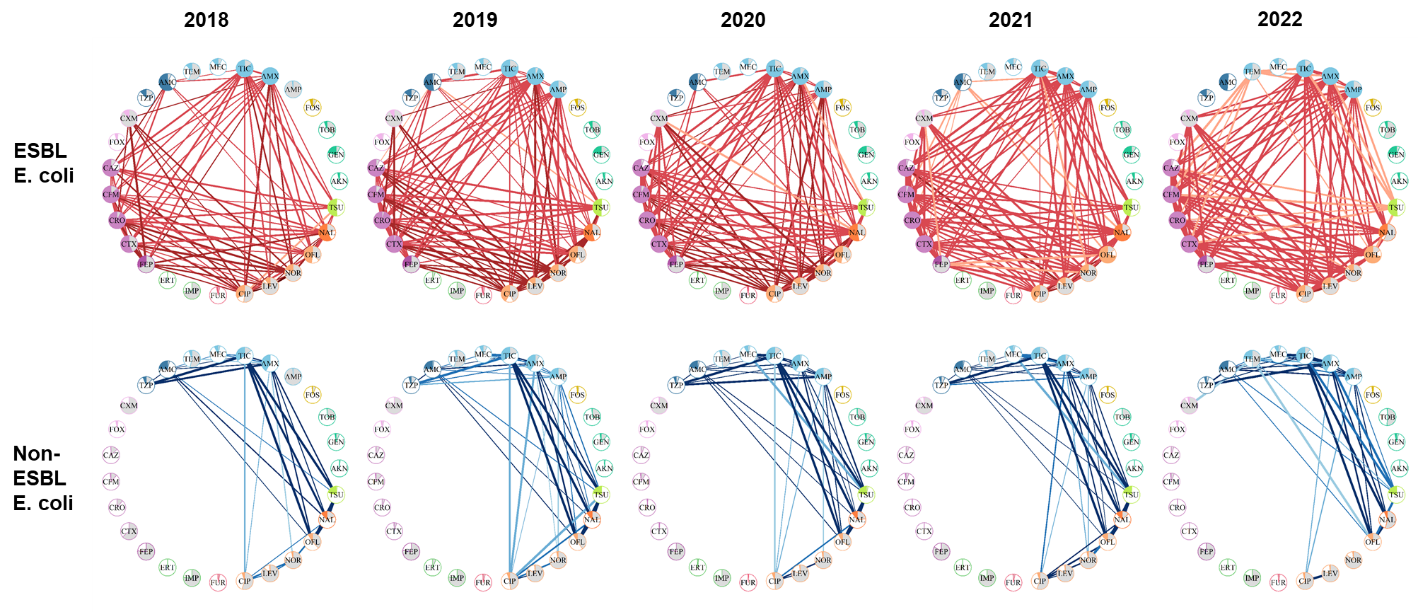

B. Results of analyses conducted with isolates coming from individuals 65 and over from 2018 to 2021

### Figure S6. Results of analyses conducted with isolates from (A) men and (B) women, from 2018 to 2021.

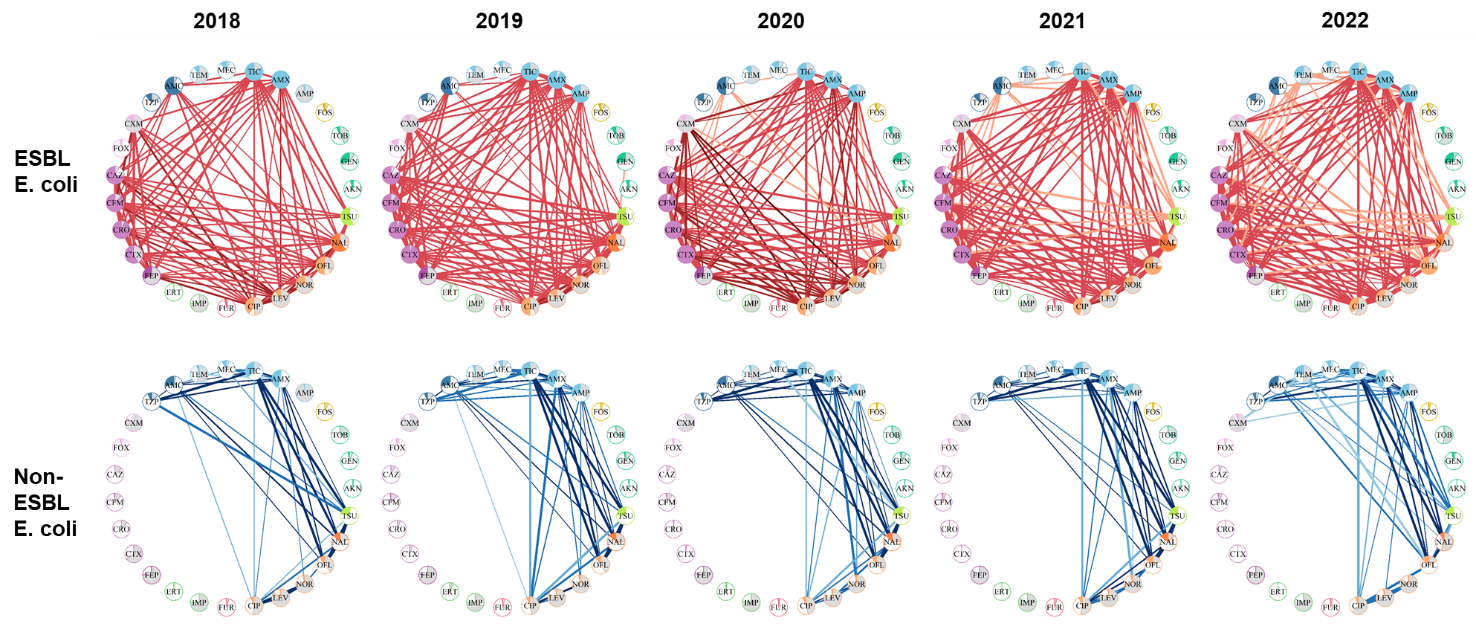

A. Results of analyses conducted with isolates coming from men from 2018 to 2021

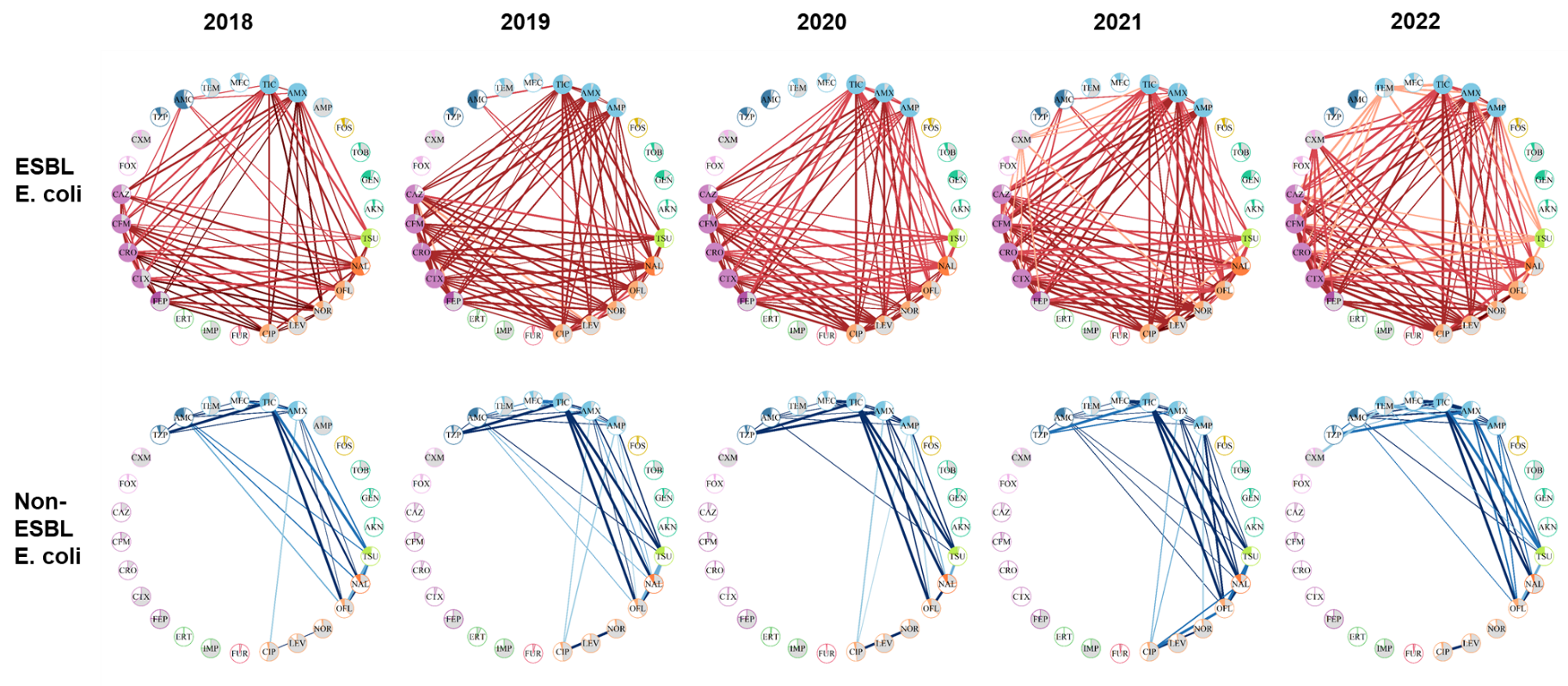

B. Results of analyses conducted with isolates coming from women from 2018 to 2021

### Figure S7. Results of sensitivity analyses accounting for the varying contribution of each region from 2019 to 2022

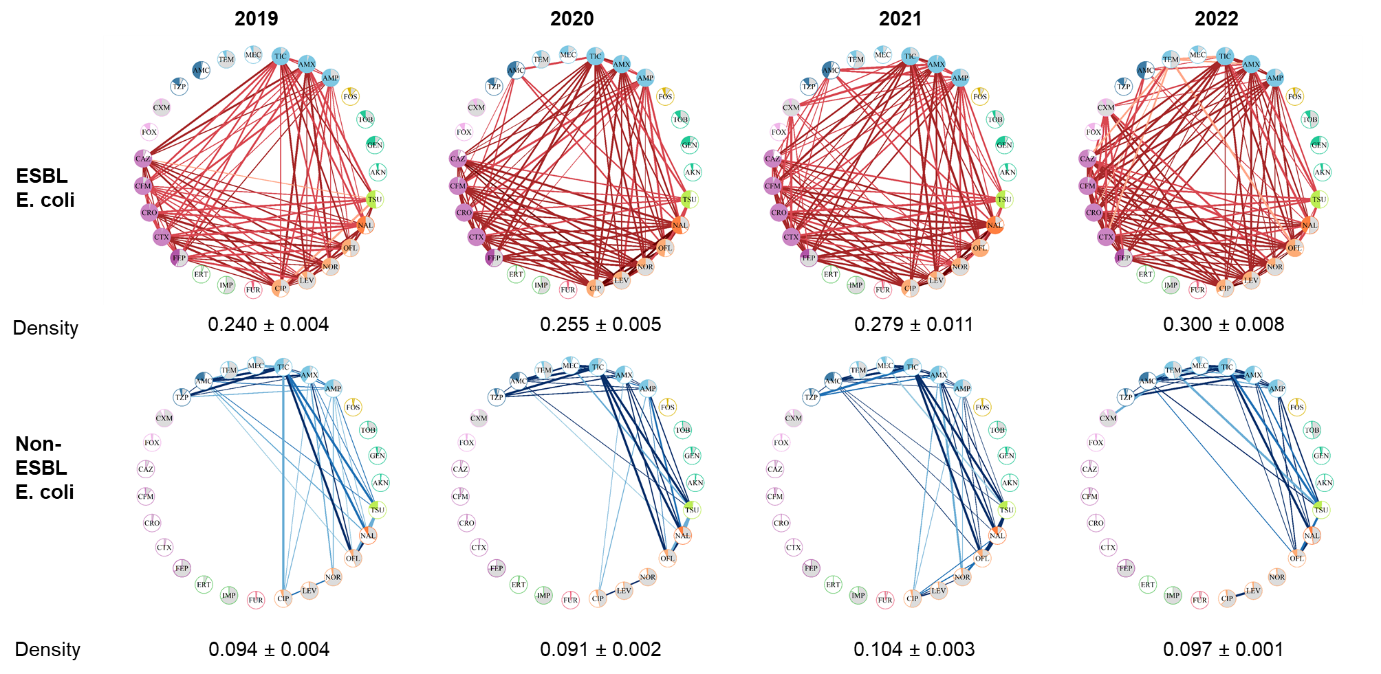
